# Automated Craniofacial Biometry with 3D T2w Fetal MRI

**DOI:** 10.1101/2024.08.13.24311408

**Authors:** Jacqueline Matthew, Alena Uus, Alexia Egloff Collado, Aysha Luis, Sophie Arulkumaran, Abi Fukami-Gartner, Vanessa Kyriakopoulou, Daniel Cromb, Robert Wright, Kathleen Colford, Maria Deprez, Jana Hutter, Jonathan O’Muircheartaigh, Christina Malamateniou, Reza Razavi, Lisa Story, Jo Hajnal, Mary A. Rutherford

## Abstract

**Objectives:** Evaluating craniofacial phenotype-genotype correlations prenatally is increasingly important; however, it is subjective and challenging with 3D ultrasound. We developed an automated landmark propagation pipeline using 3D motion-corrected, slice-to-volume reconstructed (SVR) fetal MRI for craniofacial measurements.

**Methods:** A literature review and expert consensus identified 31 craniofacial biometrics for fetal MRI. An MRI atlas with defined anatomical landmarks served as a template for subject registration, auto-labelling, and biometric calculation. We assessed 108 healthy controls and 24 fetuses with Down syndrome (T21) in the third trimester (29-36 weeks gestational age, GA) to identify meaningful biometrics in T21. Reliability and reproducibility were evaluated in 10 random datasets by four observers.

**Results:** Automated labels were produced for all 132 subjects with a 0.03% placement error rate. Seven measurements, including anterior base of skull length and maxillary length, showed significant differences with large effect sizes between T21 and control groups (ANOVA, p<0.001). Manual measurements took 25-35 minutes per case, while automated extraction took approximately 5 minutes. Bland-Altman plots showed agreement within manual observer ranges except for mandibular width, which had higher variability. Extended GA growth charts (19-39 weeks), based on 280 control fetuses, were produced for future research.

**Conclusion:** This is the first automated atlas-based protocol using 3D SVR MRI for fetal craniofacial biometrics, accurately revealing morphological craniofacial differences in a T21 cohort. Future work should focus on improving measurement reliability, larger clinical cohorts, and technical advancements, to enhance prenatal care and phenotypic characterisation.

## Introduction

Comprehensive prenatal characterisation of craniofacial development remains a challenge for obstetric ultrasound due to limitations caused by fetal position, artefacts, and technical difficulties in the 2D and 3D domain. Currently, clinical imaging techniques broadly rely on subjective assessment of facial features, and in high-risk cases, quantitative linear measurements, facial angles, and indexes, have been proposed for use with prenatal ultrasound^1–4^. Prenatal detection of face, ear and neck anomalies are low during universal second-trimester ultrasound screening in pregnancy. Indeed, a recent systematic review found them to have the lowest detection rate of 46 organ groups assessed, with a sensitivity of 32.3%^5^. Prenatal detection of facial anomalies, even in high-risk groups referred for a targeted specialist ultrasound, are even more likely to remain undetected if isolated, or, if presenting with an additional non-genetic body structural anomaly^6^. The fetal MRI craniofacial clinical literature usually describes subjective imaging assessments, and are often clinical reviews or case studies, however, its complementary role to ultrasound is often highlighted. For example, a recent historical cohort study of 45 patients referred to fetal MRI for a wide range of suspected craniofacial malformations at an anomaly level, e.g. cleft lip and palate, craniosynostosis, hyper/hypotelorism, ear structure anomalies, facial cysts and masses, found MRI was more likely to make a confident diagnosis and less likely to over-diagnose when compared to ultrasound^7^.

### Imaging craniofacial development in-utero

Imaging the craniofacial complex prenatally requires expert image acquisition, precise 2D image planes, or, 3D data that can be aligned to the region of interest (ROI). A quantitative assessment, for a prenatal phenotypic characterisation, is thus time-consuming and subject to observer variation. International guidelines for routine mid-trimester ultrasound, performed at approximately 20 weeks gestational age, GA, suggests the facial examination should be limited to a qualitative assessment of the upper lip, orbits, and an optional examination of the mid-sagittal facial profile and nasal bone^8^.

The increased use of 3D data in fetal ultrasound and the expanding applications of structural fetal MRI have resulted in the feasibility of using extended biometrics methods to better characterise and/or diagnosis subtle craniofacial dysmorphology^9,10^. In an expert consensus paper, Merz et al (2012) suggested a targeted craniofacial examination to include 3D ultrasound, with multiplanar and aligned 2D views to allow the biometric assessment of the nasal bone, NB, frontomaxillar facial angle, FMA, inferior facial angle, IFA, orbital diameters, OD, interorbital distance, IOD, and outer orbital distance, (or bi-orbital distance, BOD), maxilla width, MxW, and mandibular width, MdW (all in addition to standard head biometry i.e. head circumference, HC, occipitofrontal diameter, OFD, and biparietal diameter, BPD)^1^. However, few antenatal imaging studies have sought to comprehensively assess multiple craniofacial biometric profiles, often focussing on fetal estimated weight parameters (i.e. HC, OFD, BPD, in addition to abdominal circumference, and femur length), orbital and mandibular regions^3,11–15^.

### Fetal MRI for craniofacial assessment

Toren et al in (2020), confirmed the feasibility of multiple manually extracted fetal MRI craniofacial biometrics^16^. The authors reviewed the use of eight fetal MRI 2D measurements related to the mandible and nasal cavity, these included the, previously cited measurements, IFA, BPD, and IOD, and four new measurements, the mandibular anterior-posterior diameter, mandibular vertebral length, maximum nasal length, septal height, and septal length. The authors highlighted the additional need for structural radiological biomarkers to characterise fetal facial development and, importantly, noted that 70% (843 MRI scans) had to be excluded from their final sample due to motion artefact that degraded the image quality or the absence of a true orthogonal plane to produce the required measurement.

Except for the established cranial vault measurements (HC, OFD, BPD), there are limited examples of fetal MRI craniofacial reference ranges. Noteable anatomical areas examined are the orbits^17–19^, and includes our previous work on the automated extraction of fetal 2D orbial biometry from 3D volumes^20^; the mandible^21^ and more recently maxillary sinuses^22^. A reason for slower development of the MRI craniofacial literature may be because 3D fetal MRI has focused on brain development and assessment. Indeed, the first step would be to accurately define MRI landmarks for any new measurements and to ensure that image quality enables the accurate localisation of rarely assessed structures in-vivo. Furthermore, there is also a known lack of clear consensus on formalisation of fetal MRI biometry protocols, nomogram model formulas and measurement techniques for MRI between different clinical centres^23^. T2-weighted MRI is considered the optimal choice of image contrast for fetal structural assessment due to the faster acquisition times and good fluid tissue differentiation. The effect of field strength (1.5 Tesla compared to 3 Tesla, T) on brain biometry has been shown to produce small absolute differences for some measurements. This is likely due to the increased spatial resolution at 3T and resulting in larger discrepancies particularly for smaller structures^24^. In addition, whilst brain and facial anatomical detail are diagnostically acceptable at both 1.5T and 3T, however image quality may be be better at 3T or vary according to acquisition parameters^25^. There are, of course additional sources of error due to clinician training and experience, reporting software and environment, maternal breathing and fetal motion leading to imprecise acquisition planes and calliper placement that result in increased intra- and inter-observed variability.

### Automation of biometry for fetal MRI

Motion correction methods, based on 3D slice-to-volume registration (SVR)^26^, partially resolves these challenges since the 3D reconstructed images can be reoriented to any plane. 3D-derived biometric measurements are reportedly comparable with 2D slice-wise biometry^27,28^. Yet, there may be considerable operator variability when placing landmarks in a 3D volume due to the requirement to choose the correct plane for the measurement and then define the anatomical landmarks in 3D space.

Theoretically, in addition to being faster, automation of biometry should also allow reproducible biometric measurements. Recently, there have been several proposed methods for automated fetal MRI biometry with deep learning brain measurements such as biparietal and transverse cerebellar diameters and atrial diameters^29,30^ in 2D slices and ocular measurements using registration and deep learning^20,31^ in 3D motion-corrected images. However, outside of the cranial vault, there have been no reported automated solutions for craniofacial measurements for fetal MRI.

### Contributions

In this study, we formalise the first landmark-based protocol for craniofacial biometry for 3D T2w fetal head MRI in the atlas space and develop the first automated pipeline for extraction of 31 craniofacial biometry measurements based on label propagation. The performance of the pipeline is extensively evaluated with respect to traditional direct measurements by expert observers as well as the analysis of common errors and the effects of MRI image quality and field strength. Next, the utility of the proposed biometry protocol is assessed by quantitative comparison of 108 normal control and 24 T21 subjects, characterised by well-known craniofacial dysmorphology prenatally. In addition, we generated normative craniofacial biometry growth charts from 280 control subjects from 19 to 38 weeks GA range.

## Methods

### Fetal MRI datasets and preprocessing

Participants were scanned between 2014 and 2024 at a single site (St. Thomas’ Hospital, London, UK) and all maternal participants gave written informed consent for the use of data acquired under one of eight ethically approved MRI research studies. The datasets were acquired under different research studies: The Placental Imaging Project (PIP, REC 14/LO/1169); the Intelligent Fetal Imaging and Diagnosis (iFIND, REC 14/LO/1806); the quantification of fetal growth and development with MRI study (fetal MRI, REC 07/H0707/105); the fetal CMR service at Evelina London Children’s Hospital (REC 07/H0707/105); the developing human connectome project (dHCP, REC 14/LO/1169); the early brain imaging in Down syndrome study (eBiDS, REC 19/LO/0667); the Individualised risk prediction of adverse neonatal outcome in pregnancies that deliver preterm using advanced MRI techniques and machine learning study (PRESTO: REC 21/SS/0082); and the Cardiac and Placental Imaging in Pregnancy project (CARP; REC 08/LO/1958). The inclusion criteria for case selection included: singleton pregnancy, fetal MRI stacks with full ROI coverage, acceptable quality whole head SVR output. The normal control cohort included 314 cases without reported fetal or maternal anomalies with moderate to excellent image quality from four different acquisition protocols, from 19 to 39 weeks GA. The T21 cohort was curated primarily based on the availability of datasets with research consent and acceptable 3D head SVR reconstruction quality. In total, we selected 24 T21 cases from 3 different acquisition protocols and 29 - 36 weeks GA range.

#### MRI acquisition protocols

The included datasets were acquired with different MRI acquisition protocols depending on the recruiting study:

- 4 DS and 34 healthy control subjects scanned on a 1.5T Philips Ingenia MRI system using 28-channel torso coil with TE=80ms, 1.25×1.25mm resolution, 2.5mm slice thickness, −1.25mm gap and 9-11 stacks (iFIND, FCMR studies);
- 17 DS and 106 healthy control subjects were scanned on 3T Philips Achieva MRI system using a 32-channel cardiac coil with TE=180ms, 1.25 × 1.25mm resolution, 2.5mm slice thickness, −1.5mm gap and 5-6 stacks (PIP, PRESTO, eBIDs studies);
- 3 DS and 130 healthy control subjects were scanned on 3T Philips Achieva MRI system with a 32-channel cardiac coil using a dedicated dHCP fetal acquisition protocol with TE=250ms, 1.1 x 1.1mm resolution, 2.2mm slice thickness, −1.1mm gap and 6 stacks (dHCP, fetal MRI studies).
- 44 healthy control subjects scanned on a 1.5T Philips Ingenia MRI system using 28-channel torso coil with TE=180ms, 1.25×1.25mm resolution, 2.5mm slice thickness, −1.25mm gap and 9-11 stacks (PIP, CARP studies).

#### 3D SVR head reconstruction

All datasets were reconstructed for the whole head using two different automated SVR methods: the dedicated SVR pipeline developed for dHCP project^32^ and the optimised automated version^33^ of the classical 3D SVR reconstruction method^34^ in SVRTK package (https://github.com/SVRTK/SVRTK and https://github.com/SVRTK/auto-proc-svrtk)^35^. The reconstructed 3D head images have 0.8 mm isotropic resolution and are reoriented to the standard radiological space (see examples in Fig. 1). In order to account for the small dimensions of some of the biometrics, we applied additional resampling of 0.5mm isotropic resolution to all 3D reconstructions prior to landmark label propagation.

**Figure 1.**
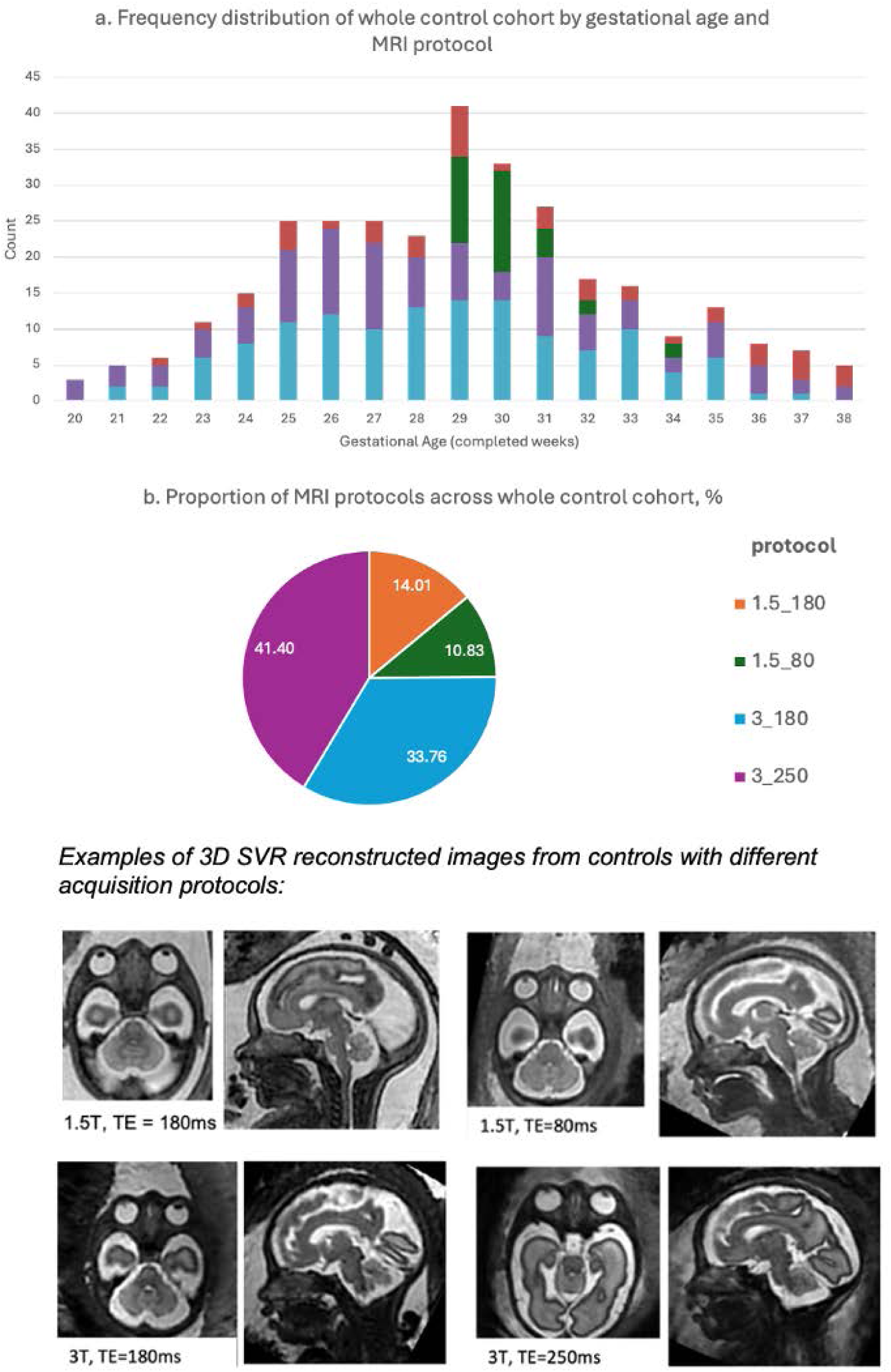
a. GA distribution of control subject datasets in the study per MRI protocol; b. proportional distribution of MRI protocols in the whole control group (MRI protocol (field strength/TE) = 1.5T/ 180ms; 1.5T/ 80ms; 3T/ 180ms; or, 3T/ 250ms); and examples of a 3D SVR fetal head reconstructions at different acquisition parameters.

The 3D whole head MRI image quality scoring protocol is shown in supplementary file S1 Fig. 8 similar to that proposed in our previous qualitative 3D MRI assessments^10^. An image score of ’good’ or ’excellent’ was given when the brain could be visualised with no, or minimal, image or reconstruction artefacts, in addition, the mid and lower facial region and the facial profile should be included within the image volume.

### Formalisation of 3D MRI craniofacial biometry protocol

Following an extensive literature search (JM), two 3-hour consensus workshops (JM, MR, AEC, AL, SA) were held which included an image review of proposed 3D landmarking points. A set of 31 biometric measurements (35 points) were agreed as relevant to clinical craniofacial assessment and feasible/reliable in terms of landmark visibility in 3D fetal MRI and included distance, angular and area measurements of the deep viscerocanial and cranial vault regions, see Fig. 2 for a visual representation of the 3D point cloud.

**Figure 2.**
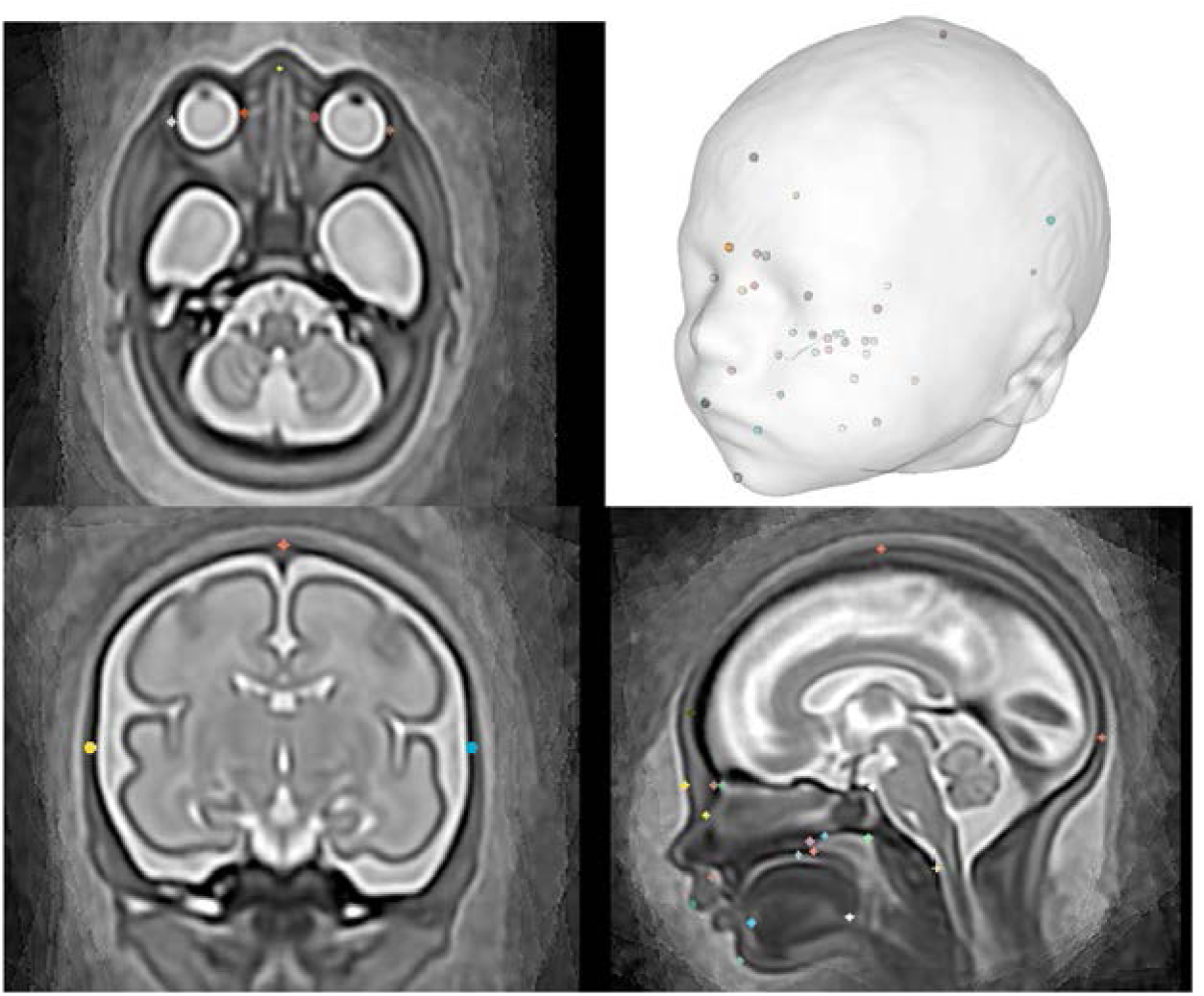
Visual representation of 3D landmarks placed within a 3D population-averaged MRI atlas (31 weeks GA)

The formalised biometry protocol, with abbreviations used in this work, is summarised in table 1 along with citations of publications that directly relate to the proposed measurement technique and/or the related reference charts^16–18,21,27,36–48^. Additionally, the wider literature supported the rationale for landmark definition and included: prenatal craniofacial biometry and anatomical MRI literature^49,50^; 2nd and 3rd trimester prenatal US measurement literature^51–53^; ex-vivo/ post-mortem anatomical studies^54,55^; and, neonatal, paediatric or adult clinical imaging literature, where relevant^22,56–59^. The label definition and location of anatomical points of interest were agreed upon in the consensus workshop, resulting in 35 points related to the measurements and a further 15 anatomical points of interest that may be of future interest (the latter not investigated in this work). These points are summarised in supplementary file S2 table 2 and 3. The points were manually placed in 3D space on to a population-average atlas of the volumetric whole fetal head^60^ using a 5mm 3D isotropic spherical ’paintbrush’ with the ITK-SNAP software(http://www.itksnap.org/pmwiki/pmwiki.php); a visual overview of the points can be seen in Fig. 2. The atlas template, extended 3D segmentation file (50 anatomical points) and the landmarking protocol are publicly available online at the KCL CDB data repository (https://gin.g-node.org/kcl_cdb/craniofacial_fetal_mri_atlas).

**Table 1.**
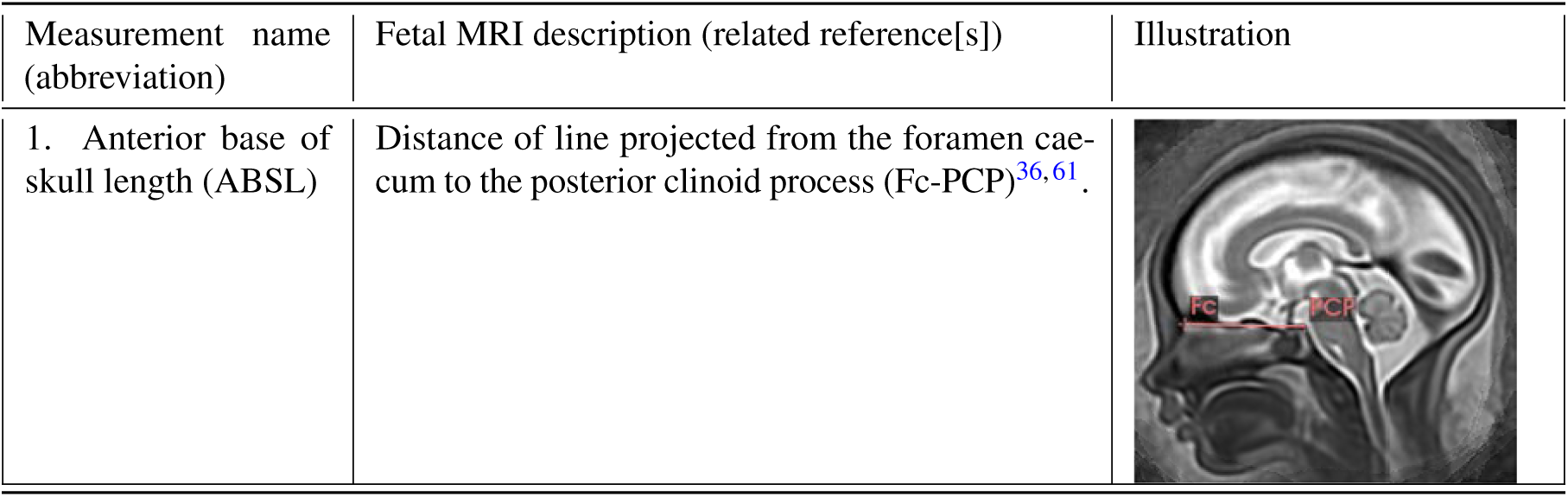
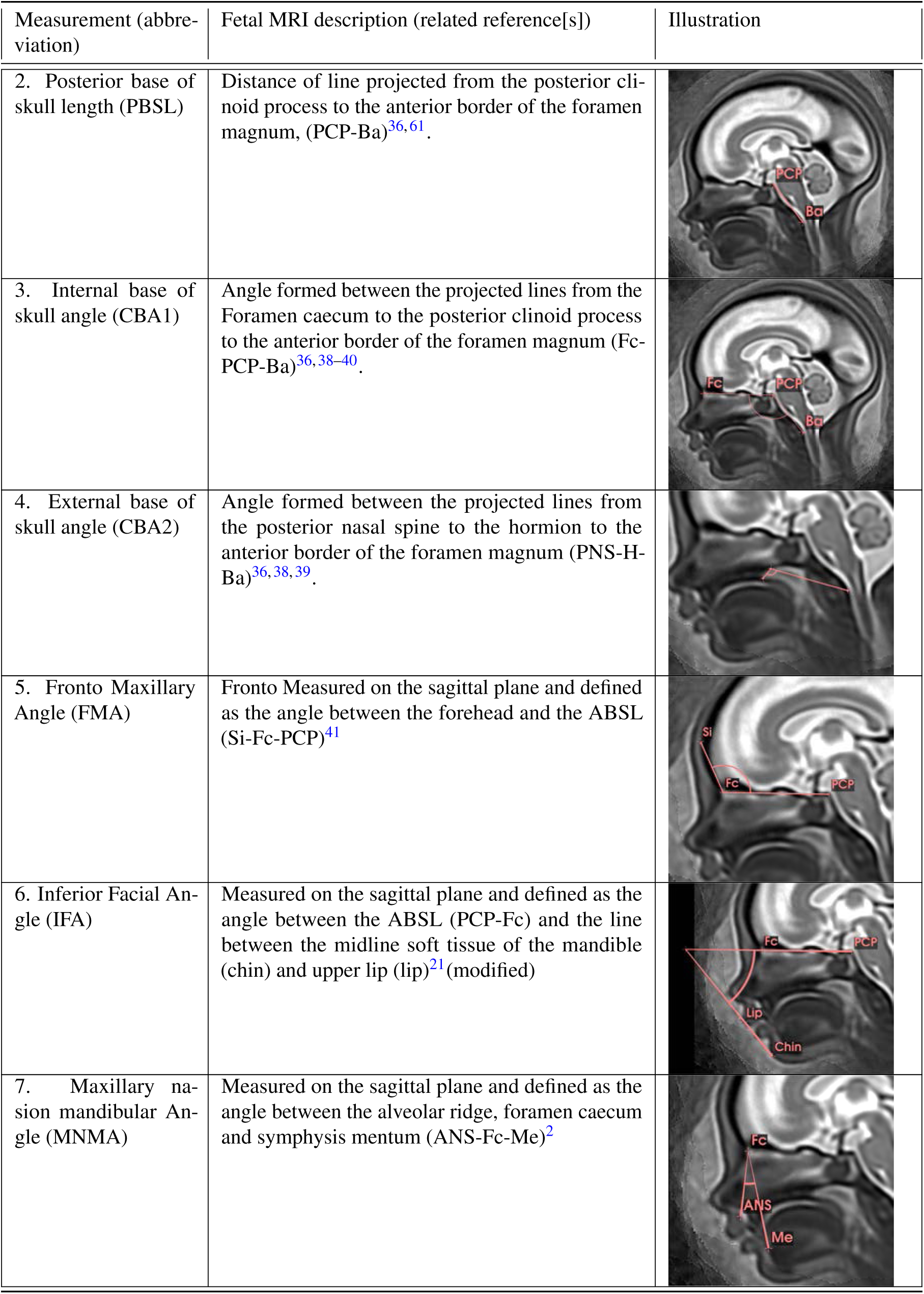
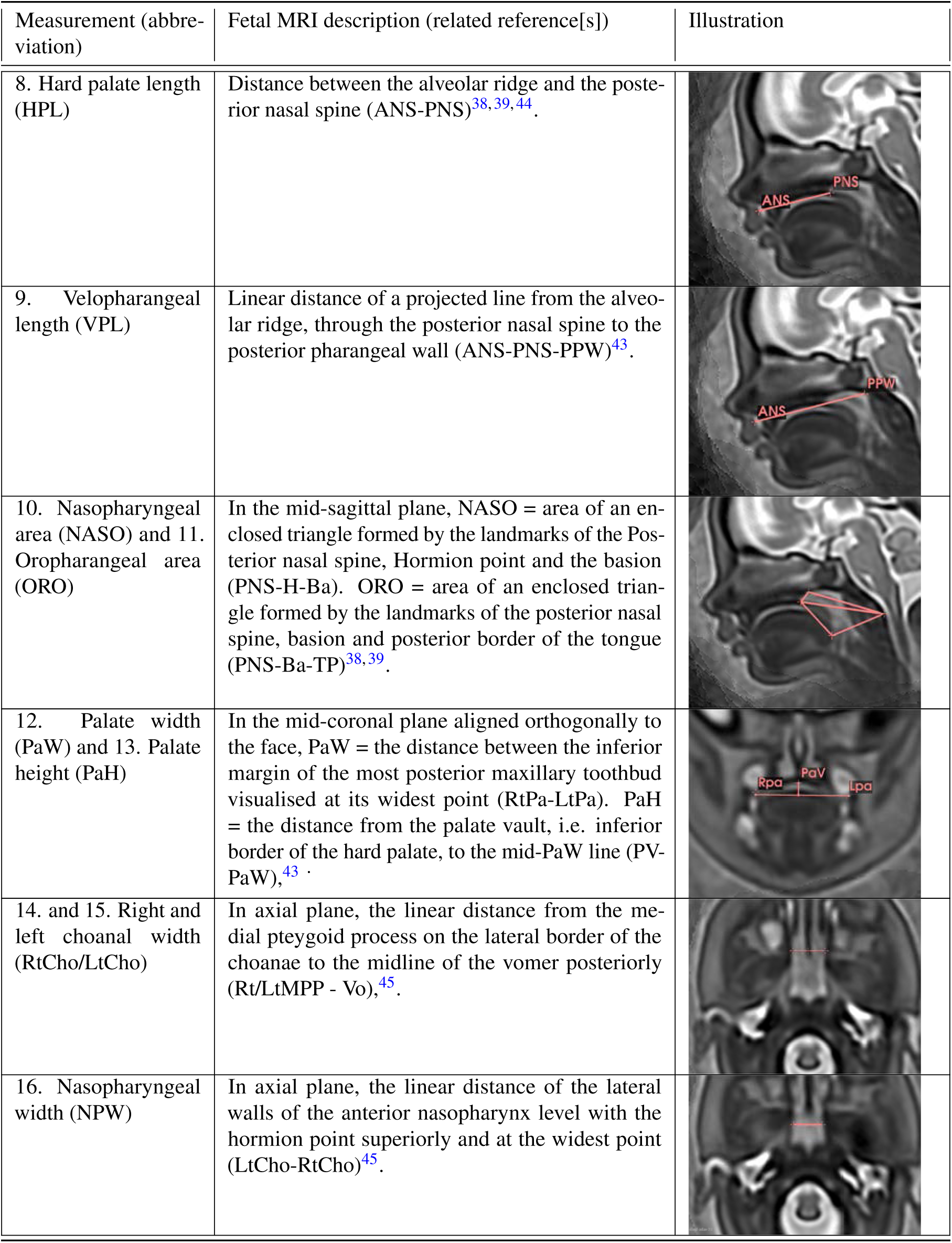
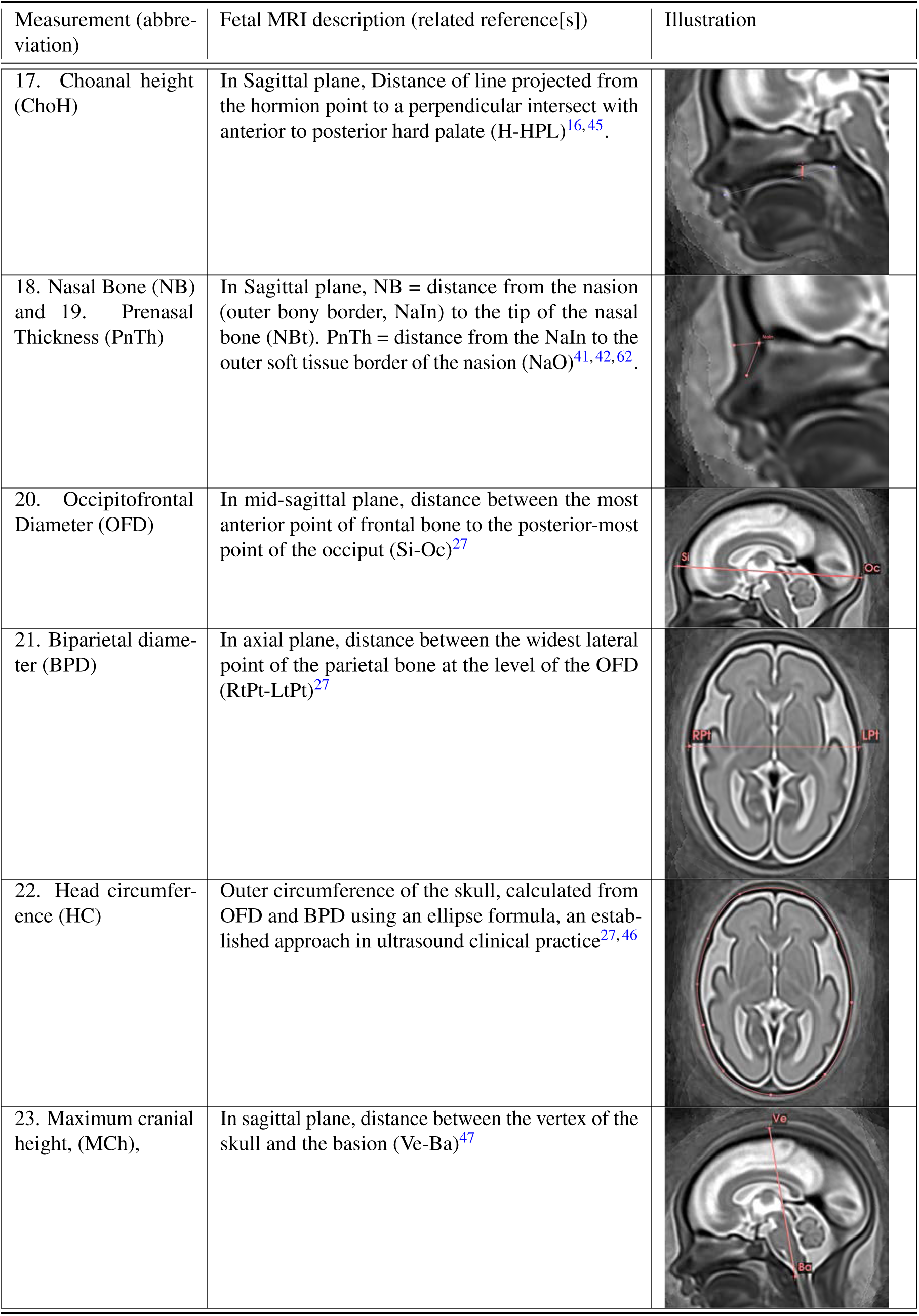
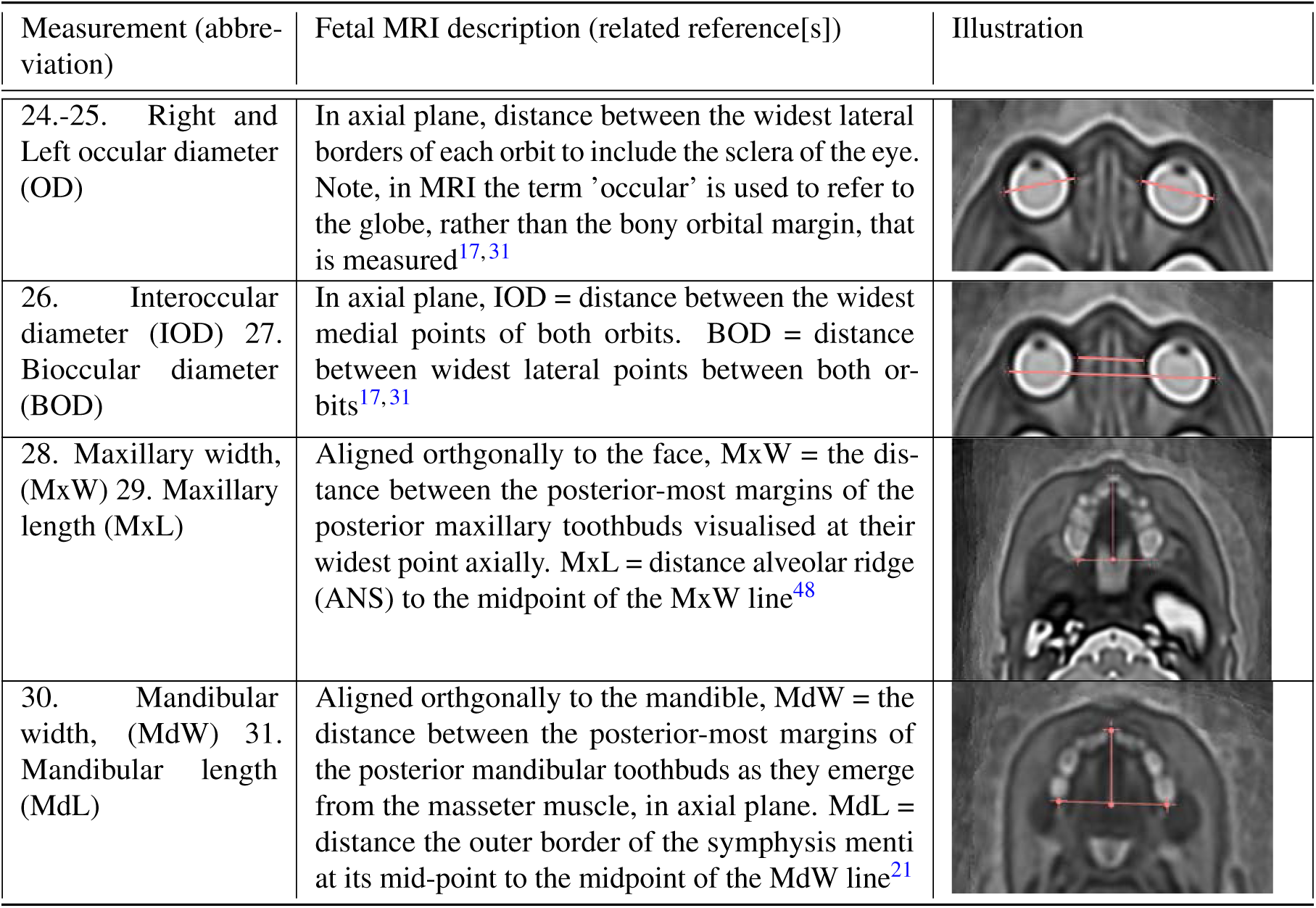
Formalised measurement definitions for the proposed landmark-based craniofacial biometry protocol with 3D T2w fetal MRI, with measurement and landmark abbreviations.

### Automated 3D craniofacial biometry pipeline

The proposed pipeline for automated biometry is outlined in Fig. 3. Firstly, the defined 3D labels from the atlas were propagated to the subject space via registration. We employed classical affine + non-rigid free form deformation registration^63^ with local normalised cross-correlation similarity metric with a 6 mm control point spacing implemented in MIRTK (https://github.com/BioMedIA/MIRTK). This registration-based approach was feasible since the 3D head images were reoriented in the standard atlas radiological space apriori after SVR reconstruction. The registration parameter were optimised for this particular task. Next, the landmark labels from the atlas were transformed to the subject space using output transformations. Label propagation was followed by computation of the defined biometry parameters based on the landmark coordinates including distances, angles and areas. Next, the landmark coordinates were computed as centre-points of propagated landmark labels in 3D world space. Lastly, this was followed by the calculation of the defined biometry measurements (table 1) using the formulas given below:

- the distance *d* between two landmarks *P*_1_(*x*_1_, *y*_1_, *z*_1_) and *P*_2_(*x*_2_, *y*_2_, *z*_2_) in 3D space:

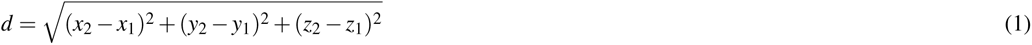
- the angle *θ* between two lines defined by landmarks *P*_1_(*x*_1_, *y*_1_, *z*_1_), *P*_2_(*x*_2_, *y*_2_, *z*_2_), and *Q*_1_(*x*_3_, *y*_3_, *z*_3_), *Q*_2_(*x*_4_, *y*_4_, *z*_4_) in 3D space:

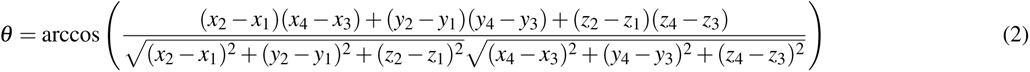
- the distance *d* between a landmark *P*(*x*_3_, *y*_3_, *z*_3_) and the center point of a line defined by landmarks *Q*_1_(*x*_1_, *y*_1_, *z*_1_) and *Q*_2_(*x*_2_, *y*_2_, *z*_2_) in 3D space:

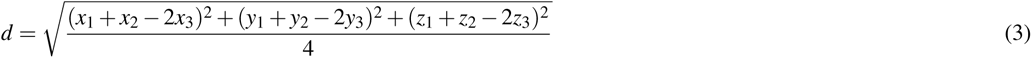
- the area *A* of a triangle from three landmarks *P*_1_(*x*_1_, *y*_1_, *z*_1_), *P*_2_(*x*_2_, *y*_2_, *z*_2_), and *P*_3_(*x*_3_, *y*_3_, *z*_3_) in 3D space:

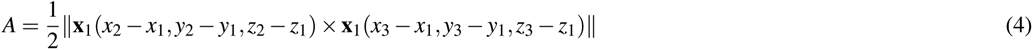
- the length of an ellipse L is calculated by a formula using two perpendicular lines (the OFD and BPD), each defined by landmarks *P*_1_(*x*_1_, *y*_1_, *z*_1_), *P*_2_(*x*_2_, *y*_2_, *z*_2_), and *Q*_1_(*x*_3_, *y*_3_, *z*_3_), *Q*_2_(*x*_4_, *y*_4_, *z*_4_) in 3D space:

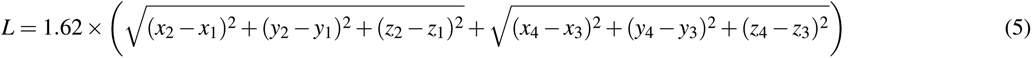

The implemented function for landmark-based biometry calculations *craniofacial-biometry* is publicly available as a part of SVRTK package.

**Figure 3.**
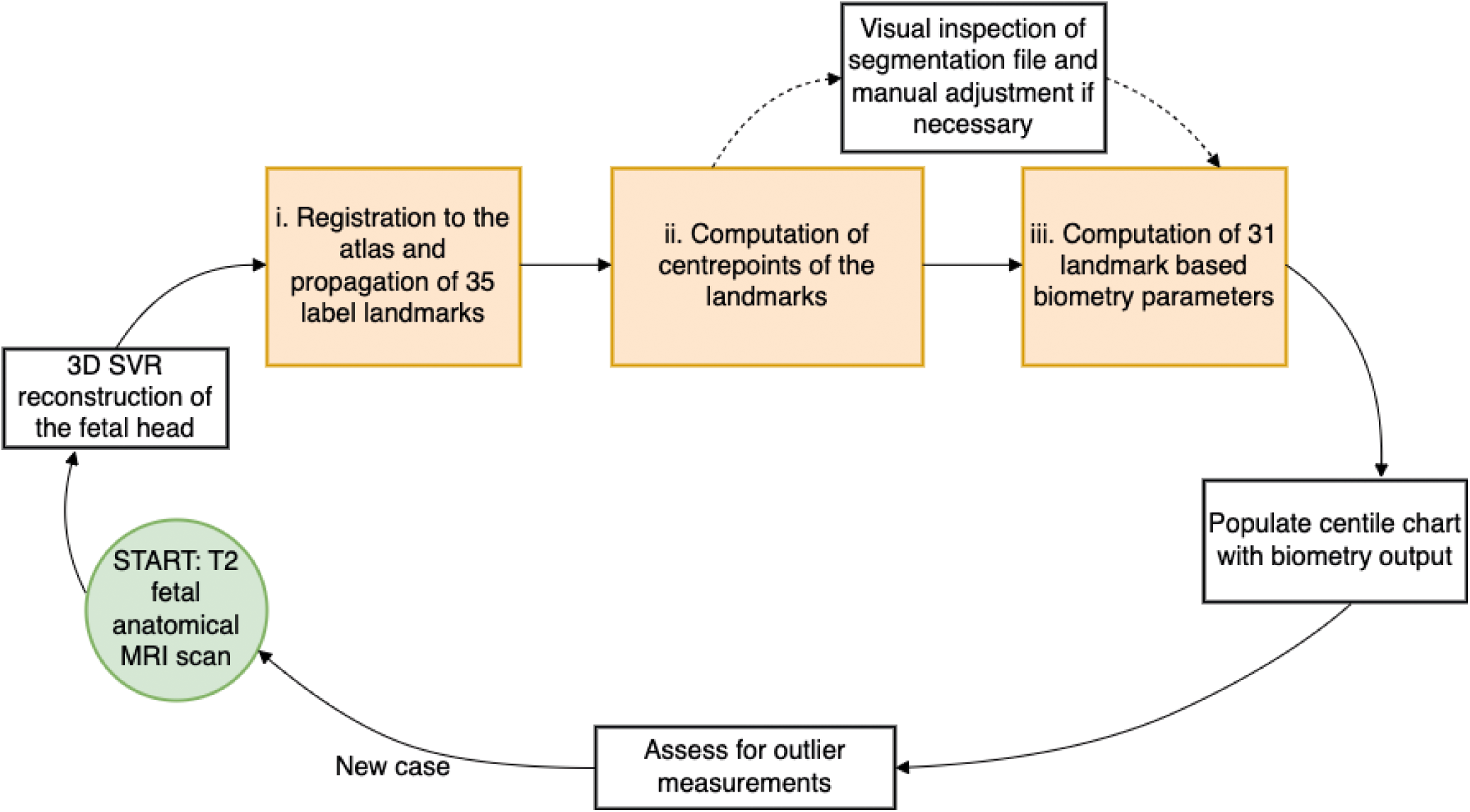
Proposed pipeline for atlas-based 3D craniofacial biometry for fetal MRI (orange boxes)

### Evaluation of the proposed biometry protocol

With the pipeline outputs of the automated label segementation files and 31 biometrics per subject, we performed an extensive evaluation of the feasibility of the proposed protocol and pipeline on normal and T21 cohorts from different acquisition protocols. This included qualitative assessment of landmark localisation in 132 datasets followed by extensive quantitative evaluation based on comparison with manual measurements in 10 cases.

#### Qualitative visual evaluation of landmarks

A single operator visually inspected all automated 3D points for 132 subjects (later used in normal and T21 comparison). The whole head SVR volume and landmarks for each subject were loaded into Slicer 3D in turn to detect any major errors in the landmark placement. 35 landmarks per subject related to biometry were inspected and scored as ’minimal or no’ error, or as having a ’major’ error (i.e. requiring a manual change of more than a few millimeters, degrees or *mm*^3^). Any measurement that was documented as an outlier (based on the distribution of cohort), had a detailed inspection and qualitative comments on the suitability of the related landmark placement. During this assessment each case was rescored for image quality, blinded to the intial image score during data curation and including an additional criteria to those presented in supplementary file S1 Fig. 8 so that *’reasonable image contrast to differentiate deep naso-oral soft tissue structures’* was also considered in the qualitative assessment.

#### Measurement Validation, intraobserver, interoberver, intramethod and intermethod

We performed assessment of intra-, inter-observer and intermethod variability in a subset of 10 random cases (from the control and T21 cohorts) given in supplementary file S2 table 4. Intraobserver repeatability was assessed by repeating measurements on the subset using a similar method to the label propagation pipeline^64^. That is, the point related to the biometric was placed using ITKsnap software, with a 5mm 3D sphere, and then the biometry was indirectly calculated automatically from the centroid centre points. The assessment was repeated after a washout period of 2 weeks, to reduce reviewer bias.

To understand the expected variability within clinical raters, the interobserver reproducibility was assessed with three fetal radiology experts. After importing the SVR volume into MITK workbench, the use of coupled cross-hairs in the x-y-z planes allowed for fine and precise adjustment of image planes within the 3D volume, required for the direct calliper measurements.

The absolute and relative differences were calculated for intra-observer, inter-observer and the automated measurements compared with the most experienced fetal radiologist, considered to be the ground truth. Bland Altman plots were constructed to visualise the variability of the automated and manual observers and compared to the most expert observer. An intraclass correlation coefficient (ICC) (two-way random effects model with absolute agreement) was performed to statistically assess the reliability of both systematic and random error to include; intraobserver (single observer, repeated indirect measurements), interobserver (3 expert raters, single direct measurements), and intermethod agreement (4 human observers and automated measures, single measurements). The ICC interpretation threshold values were reported as: *<*0.50 = poor; 0.50 - 0.75 = moderate; 0.75 - 0.90 = good; and, *>*0.90 = excellent as defined by Koo and Li (2016)^65^. A Cronbach’s Alpha test was applied to assess internal validity for each measurement, as a high internal error within the subset will reduce the power and therefore validity of the ICC^66^. A threshold of 0.7 was set as a minimum value to interpret the ICC value safely.

All human observers were asked to rate their diagnostic confidence of measuring on a binary scale (confident/not confident) for all the measurements or landmarks, where relevant. Due to the small sample size the results were presented descriptively.

### Comparison of normal and abnormal cohorts

Next, in order to assess the clinical utility of the proposed biometry pipeline for automated analysis of a large number of craniofacial biometrics, we ran a multivariate analysis of covariance (MANOVA) to assess the impact of scanner field strength with GA as a covariate (control group only). A comparison with 24 T21 cases and 108 GA-matched normal control cases, including three different acquisition protocols, was tested with a univariate analysis (ANOVA) using robust standard errors to account for multiple comparisons^67^. Posthoc power calculations were included to assess the risk of type 2 error and magnitude of effect size (using, partial estimated squares, *η*^2^)^68^. In addition, the centile normative range charts were used to assess the proportion of T21 cases falling above the 95% or below the 5% for gestation for the most relevant biometrics identified by the ANOVA.

To ensure the test assumptions were met for the analysis of variance, normality was evaluated using Q-Q plots and Shapiro-Wilk tests, multicollinearity was assessed with a Pearson’s correlation coefficient, tabulated correlation matrix, and a variance inflation factor (VIF) test was performed to detect severe multicollinearity (i.e. VIF *>*10). Lastly, linearity was visually assessed with scatterplots of the variable against GA and lastly, homogeneity of variance was assessed with a Levene’s Test,^69^. Parameter estimates using robust standard errors^67^ and using the HC3 method^70^ are reported for the analysis of variance to account for any violations in test assumptions.

### Normative craniofacial biometry growth charts

Next, the automated label propagation for all suitable control cases from 19 to 38 weeks GA was performed to create extended GA nomograms for the proposed biometry protocol. The 3D points were visually inspected and corrected if necessary, where required. After extraction of all biometric measurements, the 5th, 50th, and 95th centiles for the automated biometry results were calculated based on the widely used statistical approach for growth chart construction^71^ similar to fetal MRI brain charts described by Kyriakopoulou et al. (2017)^27^. Normative range plots with centile trendlines and quadratic formula (or linear formula where relevant) formula were then produced.

### Image Processing and Statistical Analysis Software

All image processing steps, including image reconstruction, reorientation and landmark propagation, were implemented using SVRTK and MIRTK packages.

All image review software used was open source and compatible with nifti and/or dicom format 3D image volumes. Image landmark labelling for the atlas template and for the intraobserver measurements was performed in ITKsnap. Interobserver measurements were performed in MITK workbench. The review of labels for data quality was performed in 3D slicer^72^.

Data was analysed in Excel (Microsoft Excel for Mac, Version 16.85, 2024; descriptive statistics, plots), SPSS Statistics (IBM corp, version 29.0.2.0 (20), 2023; growth curves, ICC, ANOVA, MANOVA), and, RStudio (R version 4.3.3 (2024-02-29, data visualisation and outlier assessment).

## Results

### Evaluation of reliability of the proposed biometry protocol

#### Visual assessment of the proposed automated method

The automated pipeline produced an output of labels, centre-points and biometrics for all cases. The quality of the label placement is fundamental to the centre-point extraction and therefore the subsequent biometric calculation, thus a detailed assessment of image quality and appropriateness of landmark placement is described below:

#### Visual inspection and outlier assessment of all subjects

The 132 subjects used in evaluation the had paired SVR and landmark data available for visual assessment, which took approximately 5 minutes per case. Of the 4620 total landmarks assessed, n = 15, 0.3%, were deemed as requiring major editing/unsuccessful by a single operator. The 15 unsuccessful labels were all in SVR images of moderate quality and no single subject had more than one major label error. Qualitative comments were collected about the landmark placement, with the most comments made for the lip (n = 3), posterior tongue (n = 3) and chin (n = 2). The limitations in image quality described were related to low contrast resolution and noise in the lower facial region as well as compression of superficial soft tissue by external structures e.g. placenta or maternal uterine wall. Poor visibility was also described in the naso-oropharyngeal area, which received multiple mentions (n = 17) largely due to fluid motion artefact, especially in the choanal region. Fig. 4 gives an overview of the SVR QA results stratified by field strength over gestational age and image examples of major landmark errors.

**Figure 4.**
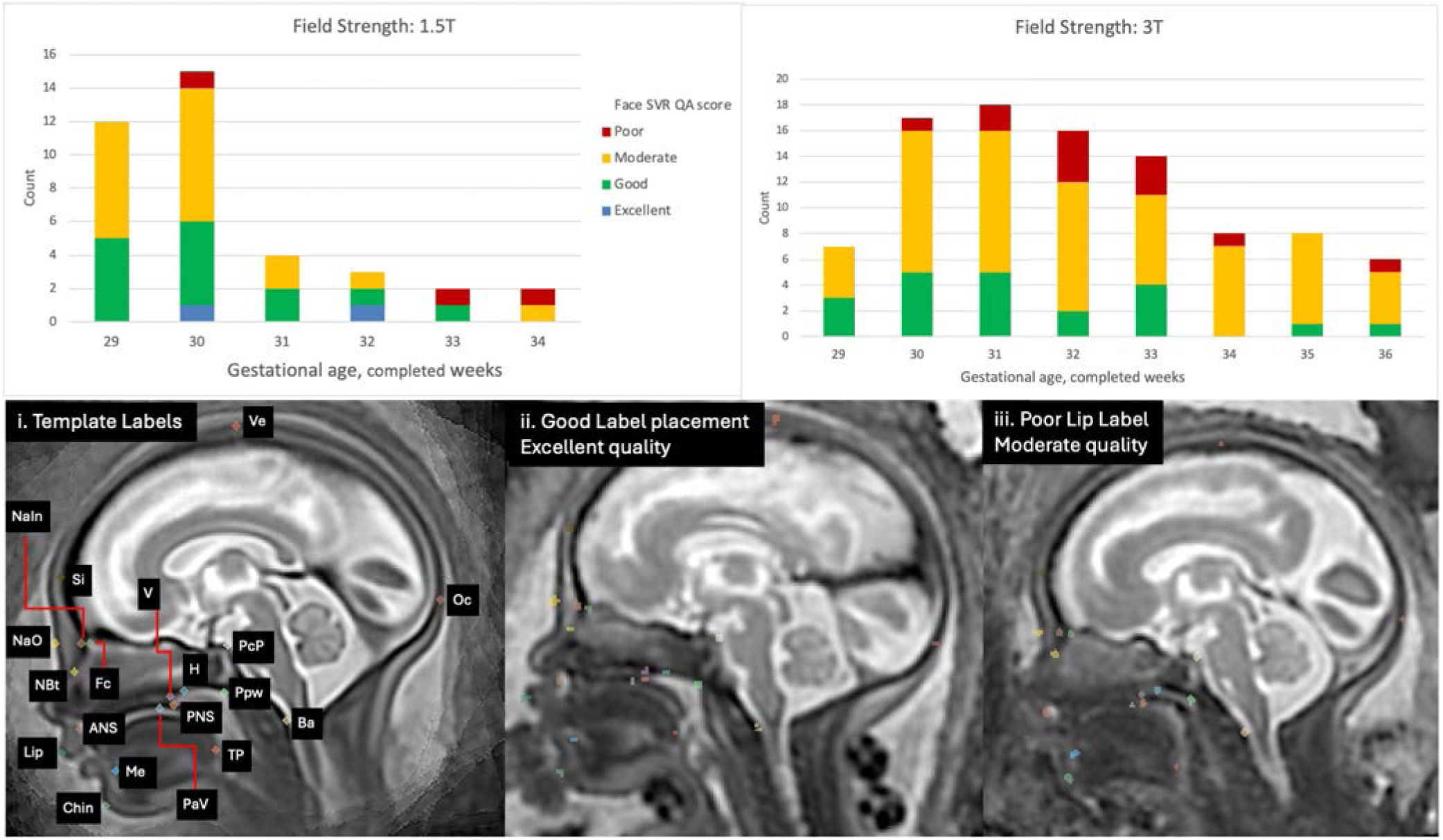
Upper row: Face SVR quality assessment results stratified by MRI field strength (1.5T/3T) and gestational age. Lower Row: i. automated landmarks with labels, ii. Example: Excellent quality SVR with good landmark placement, iii. Example: Moderate quality SVR (poor contrast resolution lower face), with poor lip label placement due to limited boundary definition adjacent to maternal tissue. (Label key: Ve=vertex; Oc=occiput; Si=sinciput; NaIn=inner nasion; NaO=outer nasion; Fc=foramen caecum; NBt=nasal bone tip; v=vomer; H=hormion; PcP=posterior clinoid process; Ppw=posterior pharyngeal wall; Pav=palate vault; Ba=basion; ANS=anterior nasal spine; PNS; posterior nasal spine; Tp=posterior tongue; Me=bony mentum; Lip=lip; Chin=chin)

Outlier measurements in the control and T21 groups were defined by assessing the measurement distributions, see boxplots in supplementary file S1 Fig. 9. 57 measurements were outliers from 32/132 unique subjects, 4 (12.5%) of the 32 outlier subjects were scanned at 1.5T and the remaining at 3T MRI field strength. The image quality of the outlier subjects ranged from poor to excellent, with a moderate score having the highest frequency. Only one measurement, the internal cranial base angle, in a 33 week old fetus scanned at 3T, was an ’extreme’ outlier, i.e., it had values above the 3rd quartile and also was more than 3 times the interquartile range. The labels related to the angle measurement appeared correctly placed on visual inspection.

#### Automated and human observer agreement and reliability

The observer variability was assessed to understand if the measurement error found for the automated biometrics were within the limits expected for expert observers. 10 cases were randomly selected for review and are presented in supplementary file S2 table 4, and were balanced interms of fetal sex, MRI field strength and were not selected for quality apriori. The repeated measures (for intraobserver agreement and reliability) were performed by a single observer, a clinical researcher with 15 years fetal imaging experience. The reproducibility measures (for interobserver agreement and reliability) were performed by 3 consultant fetal MRI neuroradiologists, with more than 10 year experience each.

For the assessment of variation of the automated measurements, the most experienced consultant radiologist was considered the ground truth, to which the absolute and relative difference were compared and presented with its mean bias and limits of agreement in a selection of Bland Altman (BA) plots, see Fig. 5 (absolute and relative difference tables for all variables can be found in supplementary file S1 Fig. 10). Intermethod agreement and reliability included all observations except the first set of intraobserver measures. An acceptable relative mean bias of less than +*/−*10% was seen for 22/31 automated measurements. Despite this, all automated measurements had mean error within the range seen for human observers except for the mandibular width (mean absolute difference; automated = 5.30mm (14.62%) versus manual (range for 3 observers) = −0.99 - 1.28mm (−2.63 - 4.55%)). The automated random error in all 31 measurements was within the limits of agreement for that seen in the manual observers, however, nine measures had an unacceptably high random error of more than +*/−*20% (Cho L, PnTh, CBA2, MNMA, IFA, PAH, ChoH, NASO and ORO)^73^.

**Figure 5.**
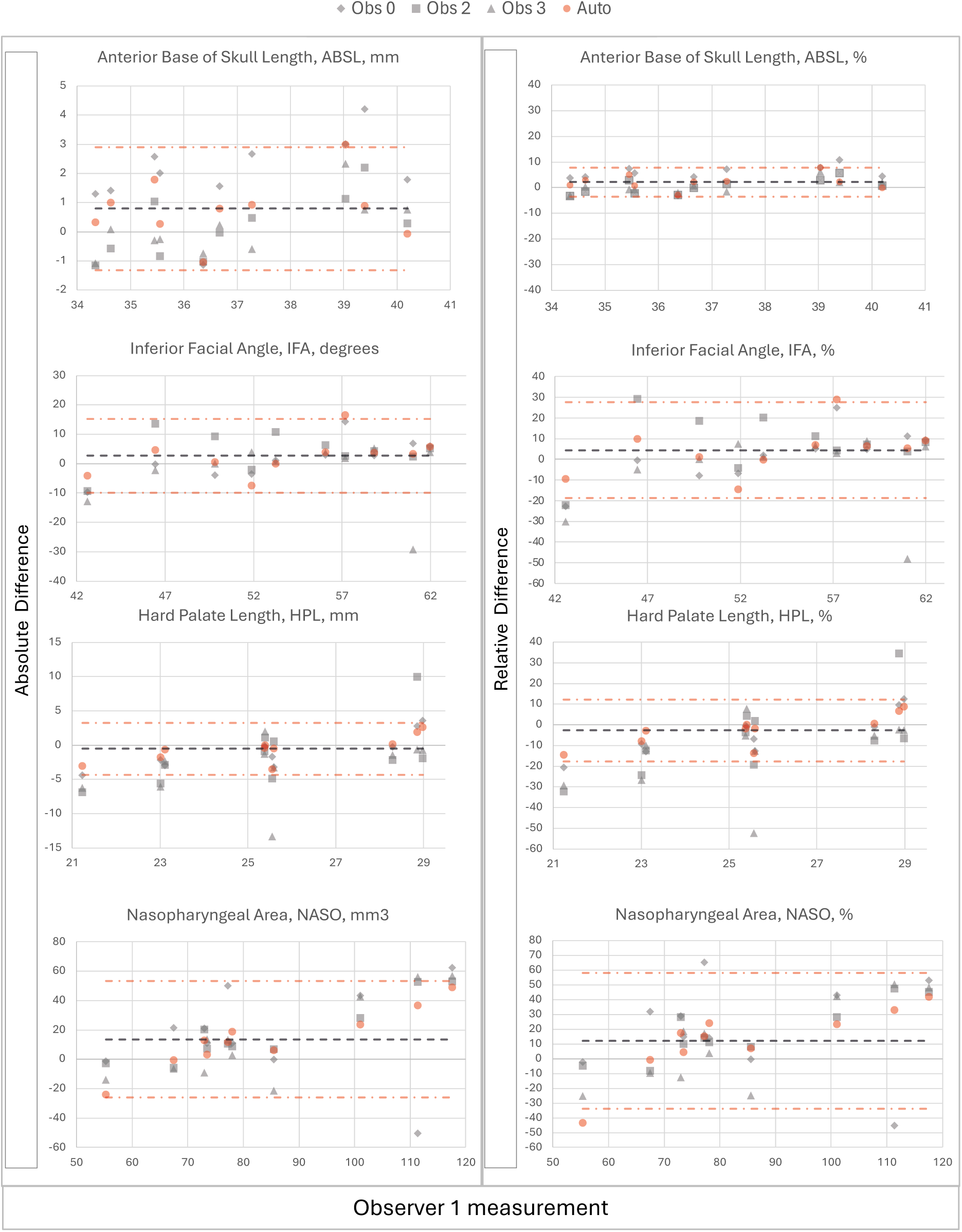
Bland Altman plots of absolute and relative difference for observer 0, grey diamond (using manual landmark-based indirect measurement method), observers 2 and 3, grey square and triangle respectively (using direct manual measurement method), and automated biometry, red circle, all compared to expert observer 1 (direct manual method) for a selection of biometrics (ABSL, IFA, HPL, and NASO). Grey dash=mean difference for automated biometry and red dash=upper and lower limits of agreement for automated method.

Intermethod reliability (i.e. for all measurements/methods including the automated) was excellent for the cranial measurements of; the anterior and posterior base of skull; the occipitofrontal diameter; biparietal diameter; maximum cranial height; internal cranial base angle; head circumference and, bi-occular diameter, (ICC range = 0.915-0.986, 95% CI range, 0.770-0.995). The intermethod reliability threshold was not below that of the interobserver reliability (expert radiologists) in any assessed variables except for the mandibular width where the ICC score changed from good (ICC = 0.904, 95% CI = 0.677-0.975) to moderate (ICC = 0.635, 95%, CI = 0.176-0.888). The ICC values were interpreted only when good internal validity of the measurement was present, i.e. Cronbach’s Alpha *>*0.70, violation of the ICC test assumptions was seen in 12/31 measurements. Full ICC results can be found in supplementary file S1 Fig. 11.

#### Human observer confidence

Subjective scoring of diagnostic confidence found that observers were least confident with measurements in the choanal and nasopharyngeal area. The intraobserver repeated measures included failed landmark placement for the nasopharyngeal space, i.e. choanal and pharyngeal width measurements and the nasal bone tip, see supplementary file S1 Fig. 12a and the average confidence scores and the variation across the three radiologist measurements are presented as boxplots in supplementary file S1 Fig. 12b. Of note, a total of 4/10 cases could not have an accurate measurement of the nasal bone performed. For the measurements obtained, the confidence scoring demonstrated that there was minimal/no variability in confidence scores, with biometrics generally scored as ’confident’. The exception was for the choanae width measurements (left, right and total width, i.e. NPW) and for cases 7 and 10 where multiple measures were more difficult. Both cases were at different scanner field strengths (1.5T/3T) and GAs (32.38 and 35.14 weeks), and image quality was subjectively scored as moderate.

### Comparison of normal control and T21 cohorts

In order to understand the clinical utility of the proposed automated pipeline, and its ability to assess differences in craniofacial development, an analysis of variance was performed on the dataset.

#### Demographics and baseline characteristics

The final retrospective sample contained 108 control and 24 T21 subjects. The mean GA for the control group was 31 weeks and 6 days (range 29 weeks and 0 days to 36 and 0 days) and for the T21 group the mean GA was 32 weeks and 3 days (range 29 and 6 days to 35 and 5 days). The distribution of MRI protocols used in the healthy control group differed from the T21 group (1.5 Tesla and echotime of 80ms in 34 (31.5%) and 4 (16.7%) respectively; 3 Tesla and echotime of 180ms in 25 (23.1%) and 17 (70.1%) respectively; and, 3 Tesla and echotime of 250ms in 49 (45.4%) and 3 (12.5%) respectively). Most datasets in the T21 cohort were performed at 3 Tesla at 180ms echotime whereas the datasets were more evenly spread in the control group, with most scans performed at 3 Tesla and 250ms. The ratio of female and male fetuses were evenly split between the cohort groups (healthy controls = 53 (49.1%)/ 54 (50.0%), and, T21 = 10 (41.7%)/10 (41.7%), the remaining fetuses had unknown sex at the time of the scan). Supplementary file S1 Fig. 13 shows the spread of fetal sex and protocols across the gestational age, with the control group having a peak at 30 weeks GA and the smaller sample of T21 fetuses being more evenly spread. The maternal ethnicity across the control group was largely of White European origin (79.6%), with 9.3% of participants reporting as having an Asian background, 3.7% as having a Black background, and the remaining participants of other or unknown ethnicity, see supplementary file S2 table 6.

#### Summary statistics of the dataset

The mean measurements and standard deviations were similar between the gestational age-matched T21 and healthy control groups, (see supplementary file S2 table 5), however, there were relatively larger mean differences between the T21 and control groups for; the occipitofrontal diameter (97.87*mm* and 101.05*mm*, respectively); inferior facial angle (55.16 and 48.97^0^, respectively); and, the nasopharyngeal and oropharyngeal areas (58.42 and 63.88*mm*^3^ and 181.41 and 173.97*mm*^3^, respectively). Box plots for the measurements indicate these differences graphically and it was noted that outlier cases were noted predominantly in the control group (see supplementary file S1 Fig. 9).

#### Growth Chart Utility and Biometric Variability Assessment

Biometric growth charts based on the age-matched control cohort were constructed for each automated biometric measurement with a calculated best-fit line and standard deviation regression equations (presented in the supplementary file S2 table 7). Using the centile ranges to assess the T21 subjects falling outside the normative ranges, the 5th to the 95th percentile were considered the threshold thus, by definition, only 10% of the healthy control cohort would be expected to fall outside of this range. 17/24 of the T21 fetal subjects had measurements that were out of range for at least one of the most meaningful 7 biometrics, i.e. the ASBL, VPL, HPL, OFD, IFA, MXL, and NASO (i.e. 71% true positive rate, TP), see Fig. 6. When considering all biometrics 23/24 cases had at least one biometrics falling outside of normative range (96% TP), with 21/24 cases having at least two measurements falling outside of the normative range (88% TP), see Fig. 14.

**Figure 6.**
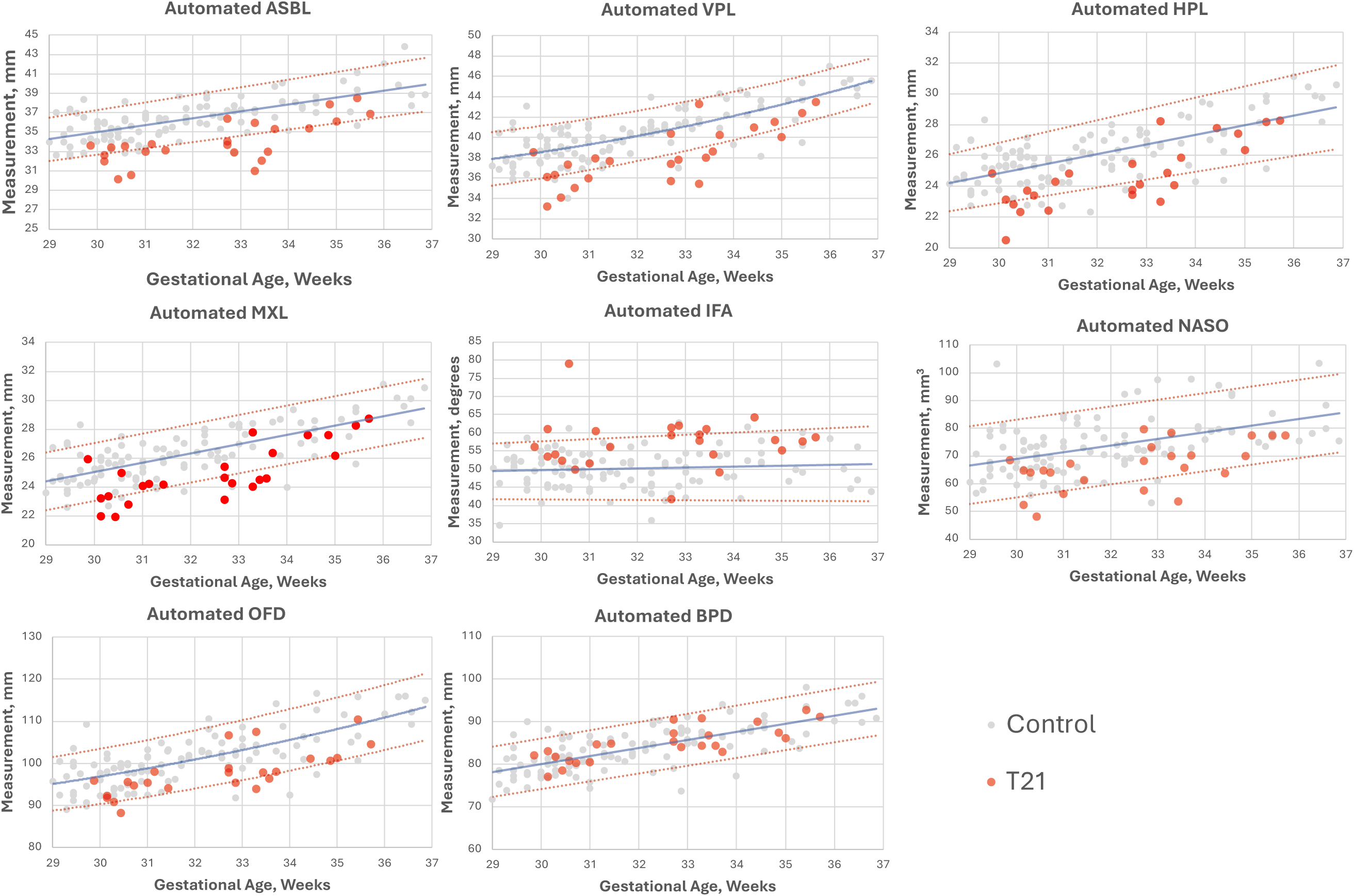
Growth charts for seven craniofacial biometrics* with statistically significant differences from control group and a large effect size, and biparietal diameter, BPD, with no significant differences between groups. (*ASBL, anterior skull base length; VPL, velopharyngeal length; HPL, hard palate length; MXL, maxillary length, IFA, inferior facial angle; NASO, nasopharyngeal area; and, occipitofontal diameter.)

### Biometric variation in the control population for subgroups of MRI field strength and GA

A MANOVA was conducted to examine the effects of MRI field strength (1.5T/3T) on the combination of 31 dependent continuous variables, i.e. the craniofacial measurements, with gestational age in completed weeks included as a covariate.

#### Variation with Gestational age

Evaluating the control group, the overall biometrics varied with GA, *p <*0.001, 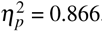, observed power = 1.00 and on univariate analysis there was no statistically significant measurement variation seen with GA for the FMA, IFA, CBA1 or ChoH (p = 0.943, 0.432, 0.639, and 0.827 respectively).

#### Variation with field strength

Within the control group there was a statistically significant difference in overall biometric variation between 1.5T and 3T MRI scans (with GA as a covariate), *p* = 0.006, 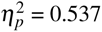, observed power = 0.99 (corrected for multiple comparisons).

### Overall biometric variation between control and T21 groups

A two-way MANOVA was performed to examine whether the dependent variables (the craniofacial measurements) differed by disease status i.e. control group or confirmed T21. The combined dependent variables of the main effect variable (disease status) showed statistically significant differences between groups *p <* 0.001, 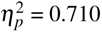, observed power = 1.000.

### Individual biometric variation between control and T21 groups

The ANOVA indicated that 7/31 variables; ASBL, HPL, VPL, OFD; IFA, MXL, and, NASO, were statistically significantly different (p *<*0.05) and had large effect sizes (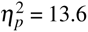 and 30.4%), with the ASBL having the largest effect, see supplementary file S2 table 8. 13 additional variables were statistically significant (PSBL, Cho_L, Cho_R, NPW, PnTh, OD_L, OD_R, IOD, BOD, MXW, MDL, MDL, and, HC) however had relatively small effect sizes of between 3.4 and 11.7%.

*Sensitivity analysis:* All non-significant variables (11/31) had a power of less than *<* 80% (PAH, PAW, CHOH, BPD, MCh, CBA1, CBA2, FMA, MNMA, ORO and interestingly NB). There were four variables reaching statistical significance despite a power below 80%; OD_R; IOD; MXW; and, MDL; all had small effect sizes of less than 5.5%. These are therefore variables that would benefit from a larger sample to meaningfully assess statistical differences in control and T21 populations with MRI.

### 2nd and 3rd trimester normative growth charts for craniofacial biometry

Following the analysis of utility of the proposed craniofacial biometry protocol for the assessment of abnormal cases we generated extended nomograms with a mixture of 3 different acquisition protocols. Datasets were included to cover the 2nd and 3rd trimester of pregnancy (*>*20 weeks GA), and with an even distribution of protocols across the GA range. Automated biometric data from 280 subjects were included, with a GA range of 19.67 to 38.62, mean 27.79 weeks. 84.3% of the sample (n = 236) was at 3T field strength and 15.7% (n = 44) was performed at 1.5T.

Linear growth patterns were seen in 5/31 measurements, IFA, FMA, CBA1, CBA2 and the NB, with the remaining variables having a quadratic growth pattern. These findings contrast with the third trimester only growth charts in section where 21/31 measurements had a linear growth pattern. In the extended GA range growth charts most variables varied with GA, however, the IFA and FMA appeared relatively stable across GA which contrasts with the four measures that were stable in the third trimester, i.e. IFA, FMA, CBA1, and ChoH. A selection of plots with the mean bestfit regression equations are shown in Fig. 7 and the full range of plots, along with dataset demographics and sample characteristics, are publicly available for research purposes online at the 3D fetal craniofacial atlas KCL repository (https://gin.g-node.org/kcl_cdb/craniofacial_fetal_mri_atlas).

**Figure 7.**
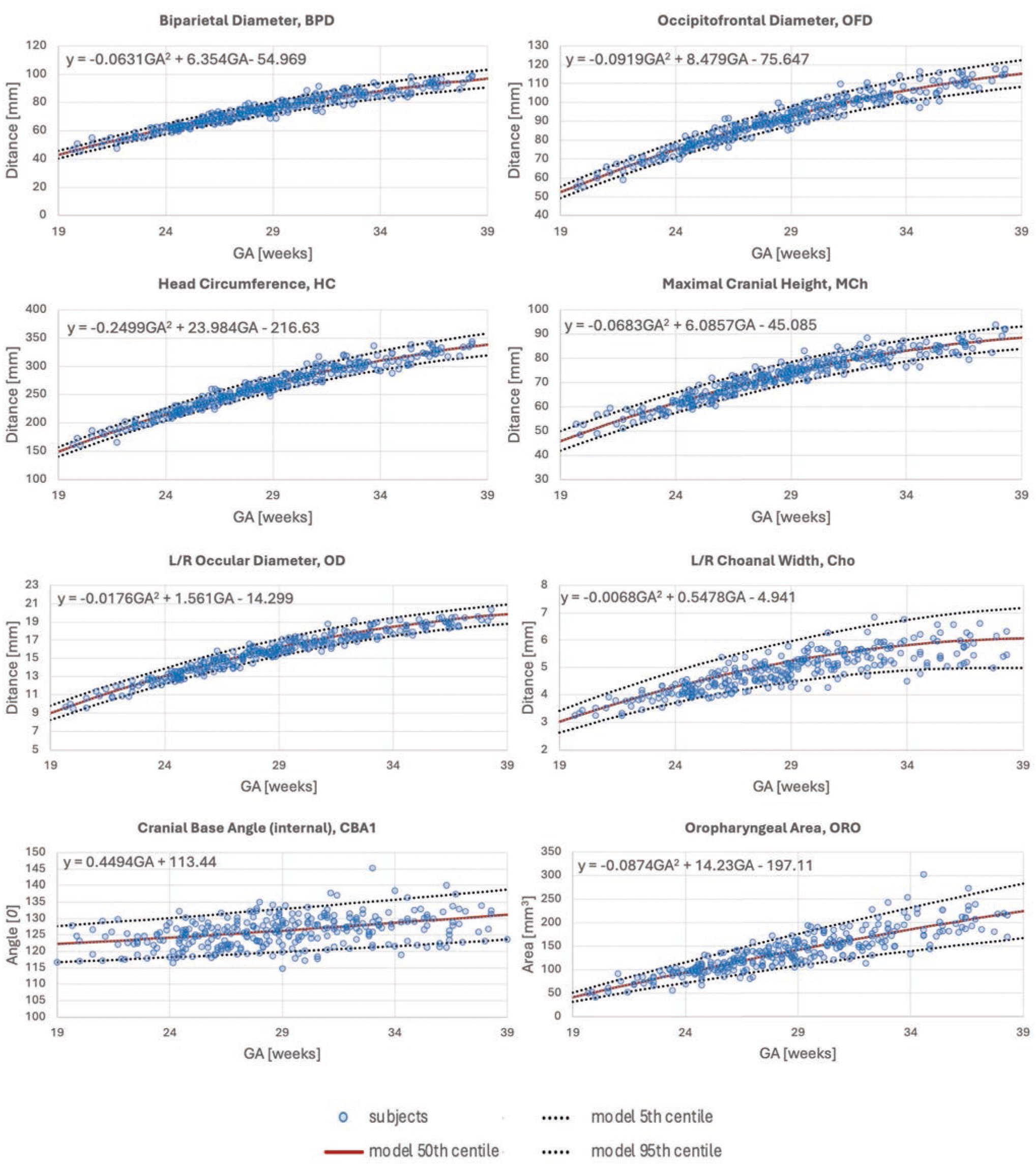
Selected growth charts for craniofacial biometry from 280 normal controls (blue circles) during 2nd and 3rd trimesters. The quadratic or linear regression equation for the 50th centile bestfit line is included on the chart (y = the mean measurement of the variable under investigation, and GA is the selected gestational age).

## Discussion

This work proposed the first comprehensive craniofacial biometry protocol and automated measurement pipeline for 3D T2w fetal MRI. We began with an extensive literature review which supported the selection of 31 biometric measurements, 29 being unique (i.e. non-bilateral), to characterise craniofacial development. The anatomical landmarks, related to the biometrics, were defined within a 3D population-averaged fetal head MRI atlas space and included 35 landmark points and corresponding mathematical 3D vector formulae for the measures. Next, we developed a pipeline for automated biometry, based on the registration of the atlas to the individual subject space and label propagation. The biometry protocol and automated pipeline were evaluated on a T21 cohort and a GA-matched healthy control cohort with a mixture of MRI acquisition protocols. This included a qualitative assessment of 132 cases by visual inspection of landmark placement, which saw a low error rate of 0.03%. The quantitative comparison included an analysis of differences between groups and an assessment of intermethod variability comparing four expert observers (manual method) and automated outputs.

This is the largest study of multiple prenatal craniofacial measurements in fetuses with confirmed T21, and whilst the MANOVA comparison of biometrics revealed statistically significant differences in many measurements (19/31), only 7 had strong effect sizes (ASBL, HPL, VPL, OFD, MXL and NASO (p*<*0.001)). These variables are all consistent with a shorter anteroposterior length of the skull and/or a smaller maxillary area, related to mid-face hypoplasia, and the findings are consistent with ultrasound and postmortem findings in T21 fetal and neonatal cohorts^59,74,75^. Interestingly the NB measurement was not found to be significantly associated with T21 in our study, which is in stark contrast to ultrasound nasal bone measurement literature^12,42^. This result is most likely due to the differing contrasts between the two imaging modalities, with MRI T2 weighted sequences having a far poorer contrast resolution of bone compared to adjacent soft tissue, affecting visibility and precision of measurements. There has been some promise of ’black bone’ imaging and, more recently, zero TE MRI sequences development to examine fetal bone, however, there are limited investigations related to craniofacial applications in fetal life^76–79^. The centiles charts related to the 7 biometrics of interest in the ANOVA model gave a true positive rate (sensitivity) of 71% that increased to 91% when all 31 variables were included. In clinical practice, the assessment of the true negative rate (specificity) is also important and not assessed here. It is highly likely that including more variables with lower effect sizes, would increase sensitivity but also increase false positives and thus reduce the specificity.

Our results indicated that the automated 3D landmark-based measurements were within the variance range of expert manual calliper placement for measurements. Systematic differences were only notable for the mandibular width and this is likely related to the manual measuring of an angular structure which requires precise slice alignment in a 3D volume of non-standard image plane, which adds complexity compared to traditional 2D measurements. It is important to consider that without a reliable ground truth measurement, the automated method produces the same results for the same datasets, if repeated, however, there are always small differences for an individuals’ repeated measures and therefore automated biometry will reduce human random and systematic error caused by differences in training, experience, environment, or fatigue. Furthermore, one of the advantages of the proposed automated pipeline is that even with manual verification (and minor refinement of landmarks) it allows significantly faster and more consistent biometry, approximately 5 minutes, in comparison to the classical manual approach that can take 25-35 minutes per case for all measurements.

Lastly, the publicly available 2nd and 3rd trimester normative biometry growth charts, based on 280 normal control subjects from 19 to 39 weeks GA, could be used as a reference for future research studies. This is a first step towards standardisation of automated 3D MRI fetal craniofacial biometry for quantitative analysis that could potentially allow efficient assessment of large cohorts.

### Limitations and future work

In this paper, we focus on the variability and reliability of fetal MRI measurements of our new baseline protocol with a first evaluation of the feasibility in a T21 cohort. Despite, yielding novel and comprehensive results in this group, a deeper analysis will be required for its application in clinical practice.

We conducted a detailed evaluation of autobiometry performance in the 3rd trimester, however further optimisation 3D landmark localisation based on deep learning, rather than atlas registration, should improve the reliability of the results. This is important to consider as the proposed method cannot exclude a landmark based on low confidence as a human observer would. Incorporation of surface-derived information could also ensure that the “upper/lower limit” measurements (e.g., skull OFD) will correspond to true anatomical values^30^. Automated quality control, QC, of 3D T2w image volumes and confidence in landmarks placement could be achieved based on deep learning classification which would require a rigorous definition of a QC protocol.

Martins et.al (2014), suggests cut off values for the limits of agreement (i.e. random errors) in measurement variability studies and their suitability for research or clinical use as +/- <5% to 10% is considered good or very good for clinical precision. However, any variability of 20-50% may still be useful for research but should be used with caution in clinical practice^73^. In our case there were large random errors observed in the manual measurements of more than 20% for nine variables and therefore any future work should exhibit caution and investigate error reduction.

Additional sources of error include the MRI scanner field strength which may impact spatial resolution, contrast resolution and the presence of artefacts. In our data, we found statistically significant differences in variability between 1.5T and 3T autobiometry. Image quality is very likely a factor that resulted in this difference with the image contrast of the lower face noted qualitatively to be a factor of variability, especially in the choanal and nasopharyngeal regions. Our analysis controlled for GA, however, there are other factors that may have also contributed to differences in the automated biometry including the impact of fetal sex or ethnicity which may all influence natural biological variation. Furthermore, taking into account the wide range of values that naturally occur at a single GA stage, a thorough assessment of the impact of biological parameters (sex, ethnicity), parental characteristics and normalisation of results to individual fetal anatomical size via measurement indexes requires further investigation.

Our constructed growth charts spanning the 2nd and 3rd trimester of pregnancy also offers opportunities for detailed characterisation of fetal craniofacial features in future clinical studies. This, together with the investigation of the growth trends in longitudinal datasets could potentially help with development of patient-specific approach for evaluation of high-risk cases. Evaluating the clinical utility of facilitating diagnoses particularly in syndromic cases is a route of future investigation. A wider range of confirmed craniofacial structural anomalies or genetic and syndromic cases with a known craniofacial phenotype could help to understand which selection biometrics are reliable for characterisation. Assessing the facial phenotype in the newborn can be made by expert geneticists and supportive imaging results that support the selection of additional diagnostic tests could be an outcome. This might also require an extension of the proposed biometry protocol with additional measurements related to the extended landmarking protocol we have provided publicly. Indeed, our MRI 3D volume landmarking protocol offers opportunities to expand on the choice of traditional biometry, to further characterise the complex craniofacial region with 3D landmark-based geometric morphometry studies or statistical shape modelling to assist diagnostic prediction, as an emerging method in clinical practice^80^.

A clinical dilemma when counselling parents includes assessing the severity of expression of the T21 phenotype, thus with advances in early volumetric imaging assessment of the fetal and neonatal brain and further understanding of the morphological covariation with craniofacial characteristics, future research could begin to explore these questions^81^.

## Conclusion

This study presents the first comprehensive craniofacial biometry protocol and automated measurement pipeline for 3D T2w fetal MRI, identifying 31 key biometric measurements, and defining corresponding landmarks in a 3D fetal head MRI atlas. An automated biometry pipeline was developed and validated with a T21 cohort and GA-matched controls, showing comparable accuracy to manual methods. Significant differences in craniofacial measurements were found in T21 fetuses, particularly highly sensitive biometrics indicating mid-face hypoplasia, a feature consistent with the T21 phenotype. The automated method reduces human error and speeds up measurements significantly, although further optimisation, especially using deep learning, is needed for clinical practice. Future work may include evaluation across earlier GA ranges and addressing potential sources of error like MRI image quality and craniofacial specific sequences at different field strengths. In addition, a focus on reducing measurement variability and exploring additional diagnostic biometrics, with growth charts and longitudinal datasets will offer a potential for personalised evaluation in high-risk cases.

## Acknowledgements

This work was supported by the NIHR clinical doctoral research fellowship to JM (NIHR300555), the European Research Council under the European Union’s Seventh Framework Programme ([FP7/ 20072013]/ERC grant agreement no. 319456) for the dHCP project; the Wellcome Trust and EPSRC IEH award [102431] for the iFIND project and the Wellcome/EPSRC Centre for Medical Engineering at King’s College London [WT 203148/Z/16/Z]; the NIH (Human Placenta Project [grant 1U01HD087202-01]) for the PIP study; NIHR Advanced Fellowship awarded to Lisa Story [NIHR30166]; the Medical Research Council grant [MR/X010007/1]. This paper represents independent research part funded by the National Institute for Health and Care Research (NIHR), the NIHR Clinical Research Facility at Guy’s and St Thomas’, and by the Maudsley Biomedical Research Centre (BRC) at South London and Maudsley NHS Foundation Trust, and King’s College London. J.O. and M.R. received support from the Medical Research Council Centre for Neurodevelopmental Disorders, King’s College London (Grant MR/N026063/1). J.O. is supported by a Sir Henry Dale Fellowship jointly funded by the Wellcome Trust and the Royal Society (Grant 206675/Z/17/Z).

## Data availability statement

Fetal MRI datasets used for this study are not publicly available due to ethics regulations. The proposed biometry protocol defined in the 3D T2w atlas space and craniofacial growth charts are publicly available online at KCL CDB fetal MRI atlas repository: https://gin.g-node.org/kcl_cdb/craniofacial_fetal_mri_atlas.

## Author contributions statement

J.M. and A.U. contributed equally to this work. J.M. performed literature review and formalised the biometry protocol, analysed the datasets, performed all evaluation experiments, interpreted the results and prepared the manuscript. A.U. reconstructed 3D SVR images, implemented automated biometry pipeline, processed all datasets and prepared all technical descriptions. A.E.C., A.L and S.A. participated in evaluation experiments and formalisation of the biometry protocol. A.F.G. participated in processing and analysis of T21 cohort cases. V.K. contributed to formalisation of the biometry protocol. D.C. and K.c. contributed to analysis of the datasets. R.W. contributed to formalisation of the craniofacial analysis pipeline. M.D. supervised various parts of the project. C.M. provided supervision and manuscript review. J.H. provided fetal MRI datasets. J.O.M. contributed to formalisation of the craniofacial analysis pipeline. C.M. supervised various parts of the project. L.S. provided fetal MRI datasets. J.V.H. and R.R. provided fetal MRI datasets and supervised various parts of the project. M.A.R. provided fetal MRI dataset, contributed to analysis of the datasets and supervised the project. All authors reviewed the final version of the manuscript.

## Additional information

### Competing interests

The authors declare that the research was conducted in the absence of any commercial or financial relationships that could be construed as a potential conflict of interest.

## Notes

### Competing Interest Statement

The authors have declared no competing interest.

### Author Declarations

The following relevant datasets were given research ethics committee (REC) approval by the Health Research Authority: The Placental Imaging Project (REC no. 14/LO/1169) – the Intelligent Fetal Imaging and Diagnosis (REC no. 14/LO/1806) – the quantification of fetal growth and development with MRI study (REC no. 07/H0707/105) – the fetal CMR service at Evelina London Children’s Hospital (REC no. 07/H0707/105) – the developing human connectome project (REC no 14/LO/1169) – the early brain imaging in Down syndrome study (REC no. 19/LO/0667) – the Individualised risk prediction of adverse neonatal outcome in pregnancies that deliver preterm using advanced MRI techniques and machine learning study (REC no. 21/SS/0082) – and the Cardiac and Placental Imaging in Pregnancy project (REC no. 08/LO/1958).

