## Supplementary Figures S1 for "Automated Craniofacial Biometry with 3D T2w Fetal MRI"

### S1: Supplementary Figures

**Figure 8.** Image examples of SVR head image quality scoring for inclusion in the dataset, scores of 3 or 4 were considered adequate quality for inclusion in the study.

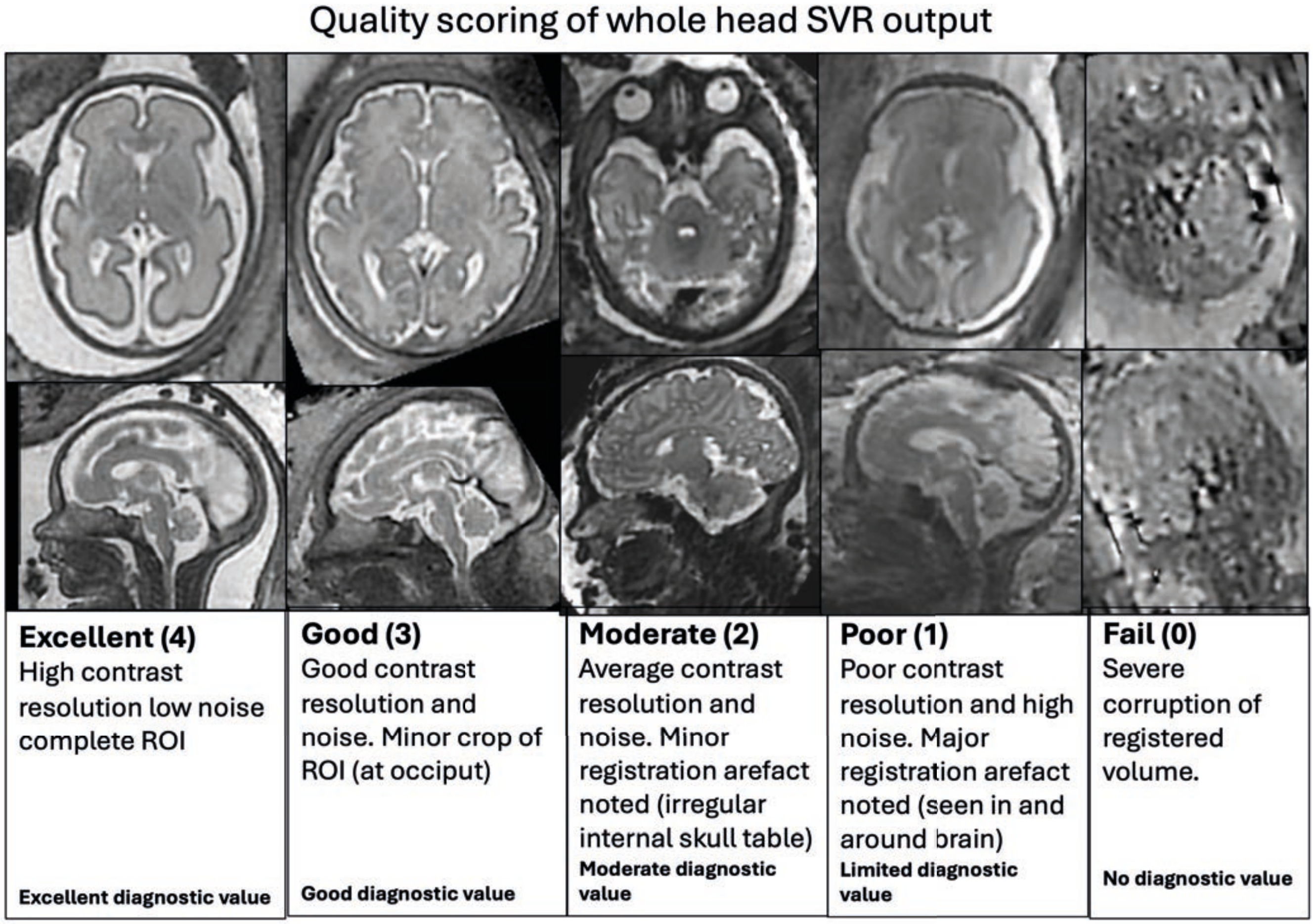

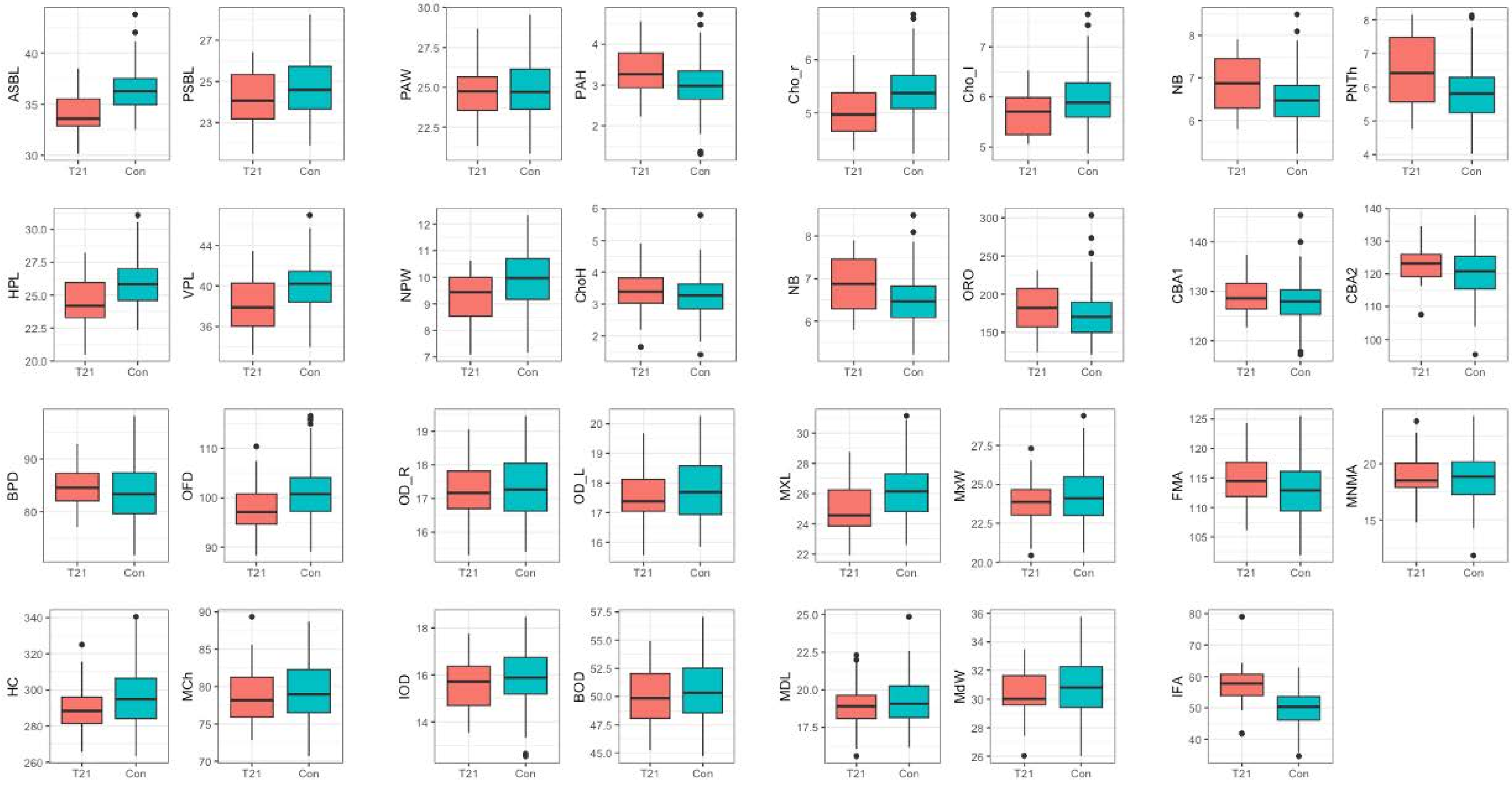


**Figure 9.** Boxplots of measurements stratified by healthy control (blue) and T21 groups (red). Central tendency is represented by the black horizontal line, and the upper and lower limits of the box represent the interquartile range (2nd to 3rd quartile). The black dots represent outliers

| Absolute difference | | Observer 0* | | | | | Observer 2 | | | | | Observer 3 | | | | | Automated | | | | |
|---|---|---|---|---|---|---|---|---|---|---|---|---|---|---|---|---|---|---|---|---|---|
| ID | Variable | N_Obs | Mean | Lower_CI | Upper_CI | abs LoA range (+/- mean) | N_Obs | Mean | Lower_CI | Upper_CI | abs LoA range (+/- mean) | N_Obs | Mean | Lower_CI | Upper_CI | abs LoA range (+/- mean) | N_Obs | Mean | Lower_CI | Upper_CI | abs LoA range (+/- mean) |
| 1 | ASBL | 10 | 1.94 | -0.80 | 4.69 | 2.75 | 10 | 0.15 | -1.99 | 2.30 | 2.14 | 10 | 0.12 | -1.81 | 2.06 | 1.93 | 10 | 0.79 | -1.32 | 2.91 | 2.12 |
| 2 | PSBL | 10 | 0.11 | -2.51 | 2.74 | 2.63 | 10 | -1.05 | -4.76 | 2.67 | 3.72 | 10 | -0.04 | -3.18 | 3.10 | 3.14 | 10 | -0.27 | -2.81 | 2.27 | 2.54 |
| 3 | HPL | 10 | -0.88 | -5.91 | 4.15 | 5.03 | 10 | -1.38 | -10.67 | 7.91 | 9.29 | 10 | -3.37 | -11.82 | 5.08 | 8.45 | 10 | -0.55 | -4.32 | 3.23 | 3.77 |
| 4 | VPL | 10 | 0.02 | -5.08 | 5.12 | 5.10 | 10 | 0.81 | -8.79 | 10.42 | 9.61 | 10 | -1.73 | -8.33 | 4.88 | 6.61 | 10 | -2.08 | -5.03 | 0.88 | 2.95 |
| 5 | PAW | 10 | -0.53 | -4.13 | 3.07 | 3.60 | 10 | -2.03 | -5.74 | 1.68 | 3.71 | 10 | -2.64 | -5.68 | 0.40 | 3.04 | 10 | -0.13 | -4.06 | 3.79 | 3.92 |
| 6 | Cho_r | 6 | 0.34 | -6.48 | 7.17 | 6.83 | 7 | -0.11 | -1.63 | 1.42 | 1.53 | 6 | 1.15 | -1.89 | 4.18 | 3.03 | 7 | 0.16 | -0.80 | 1.11 | 0.95 |
| 7 | Cho_l | 6 | -0.01 | -7.61 | 7.58 | 7.59 | 7 | -0.17 | -1.67 | 1.33 | 1.50 | 6 | 0.91 | -1.87 | 3.70 | 2.79 | 7 | -0.20 | -1.53 | 1.13 | 1.33 |
| 8 | NPW | 6 | 0.23 | -3.50 | 3.97 | 3.73 | 7 | 0.12 | -2.74 | 2.99 | 2.87 | 6 | 2.09 | -4.19 | 8.36 | 6.28 | 7 | 1.66 | -0.64 | 3.96 | 2.30 |
| 9 | NB | 7 | 0.67 | -4.20 | 5.55 | 4.88 | 8 | 2.58 | -0.71 | 5.86 | 3.29 | 9 | 0.10 | -3.88 | 4.07 | 3.98 | 9 | 1.72 | -0.26 | 3.69 | 1.97 |
| 10 | PNTh | 10 | 0.38 | -3.70 | 4.46 | 4.08 | 10 | -0.56 | -5.59 | 4.48 | 5.03 | 10 | -0.31 | -4.33 | 3.72 | 4.03 | 10 | 0.12 | -3.08 | 3.32 | 3.20 |
| 11 | OFD | 10 | 0.30 | -4.50 | 5.10 | 4.80 | 10 | -1.18 | -6.68 | 4.31 | 5.50 | 10 | 0.25 | -7.05 | 7.55 | 7.30 | 10 | 0.44 | -4.70 | 5.57 | 5.13 |
| 12 | BPD | 10 | 1.29 | -6.27 | 8.85 | 7.56 | 10 | -0.81 | -7.76 | 6.15 | 6.95 | 10 | -0.61 | -7.75 | 6.52 | 7.14 | 10 | 1.96 | -4.44 | 8.36 | 6.40 |
| 13 | MCh | 10 | 1.81 | -3.90 | 7.51 | 5.71 | 10 | 1.14 | -3.68 | 5.95 | 4.81 | 10 | 0.74 | -5.26 | 6.74 | 6.00 | 10 | -0.33 | -5.92 | 5.26 | 5.59 |
| 14 | OD_R | 10 | -0.70 | -2.95 | 1.54 | 2.24 | 10 | -0.90 | -2.58 | 0.77 | 1.67 | 10 | -1.00 | -3.26 | 1.27 | 2.26 | 10 | 0.21 | -1.60 | 2.03 | 1.81 |
| 15 | OD_L | 10 | -0.48 | -1.83 | 0.87 | 1.35 | 10 | -0.71 | -2.95 | 1.54 | 2.25 | 10 | -1.11 | -2.61 | 0.39 | 1.50 | 10 | 0.11 | -1.01 | 1.24 | 1.12 |
| 16 | IOD | 10 | 0.45 | -2.00 | 2.89 | 2.44 | 10 | -1.23 | -4.43 | 1.98 | 3.20 | 10 | 0.63 | -1.87 | 3.13 | 2.50 | 10 | 0.14 | -2.13 | 2.40 | 2.27 |
| 17 | BOD | 10 | -0.34 | -3.44 | 2.75 | 3.09 | 10 | -0.71 | -3.74 | 2.33 | 3.04 | 10 | -1.06 | -4.27 | 2.15 | 3.21 | 10 | 0.51 | -2.05 | 3.08 | 2.56 |
| 18 | MxW | 10 | 1.28 | -0.91 | 3.47 | 2.19 | 10 | 0.72 | -3.25 | 4.70 | 3.98 | 10 | -0.99 | -5.95 | 3.97 | 4.96 | 10 | 2.05 | -0.20 | 4.29 | 2.24 |
| 19 | MdW | 10 | 1.67 | -1.87 | 5.21 | 3.54 | 10 | 0.34 | -1.11 | 1.79 | 1.45 | 10 | -0.89 | -3.41 | 1.64 | 2.52 | 10 | 5.30 | 1.58 | 9.01 | 3.71 |
| 20 | CBA1 | 10 | 0.60 | -4.49 | 5.70 | 5.09 | 10 | 2.08 | -5.43 | 9.58 | 7.51 | 10 | -0.03 | -7.39 | 7.33 | 7.36 | 10 | 0.39 | -6.89 | 7.66 | 7.28 |
| 21 | CBA2 | 10 | -7.44 | -48.97 | 34.09 | 41.53 | 10 | -1.41 | -42.85 | 40.04 | 41.44 | 10 | 8.32 | -18.69 | 35.33 | 27.01 | 10 | -9.93 | -30.10 | 10.24 | 20.17 |
| 22 | FMA | 10 | 0.78 | -12.79 | 14.35 | 13.57 | 10 | 2.64 | -9.01 | 14.30 | 11.66 | 10 | -5.65 | -22.02 | 10.72 | 16.37 | 10 | 0.40 | -12.46 | 13.26 | 12.86 |
| 23 | MNMA | 10 | -10.78 | -20.54 | -1.01 | 9.76 | 10 | -3.61 | -12.23 | 5.00 | 8.61 | 10 | -10.35 | -23.02 | 2.33 | 12.67 | 10 | -7.73 | -16.16 | 0.71 | 8.43 |
| 24 | IFA | 10 | 1.65 | -11.41 | 14.70 | 13.06 | 10 | 4.27 | -8.68 | 17.21 | 12.95 | 10 | -2.53 | -23.60 | 18.54 | 21.07 | 10 | 2.68 | -9.91 | 15.26 | 12.59 |
| 25 | PAH | 10 | -1.37 | -4.07 | 1.32 | 2.70 | 10 | -1.49 | -4.09 | 1.12 | 2.60 | 10 | -0.87 | -5.08 | 3.33 | 4.21 | 10 | -0.15 | -2.40 | 2.10 | 2.25 |
| 26 | ChoH | 6 | 1.32 | -0.59 | 3.23 | 1.91 | 6 | 1.00 | -1.15 | 3.14 | 2.15 | 6 | -0.53 | -2.92 | 1.86 | 2.39 | 6 | 0.74 | -1.39 | 2.87 | 2.13 |
| 27 | MXL | 10 | -1.00 | -6.33 | 4.34 | 5.34 | 10 | 0.30 | -3.28 | 3.89 | 3.58 | 10 | 2.47 | -8.71 | 13.64 | 11.18 | 10 | -0.76 | -3.98 | 2.45 | 3.22 |
| 28 | MDL | 10 | 2.85 | -2.43 | 8.13 | 5.28 | 10 | 0.33 | -3.26 | 3.92 | 3.59 | 10 | 1.04 | -6.31 | 8.40 | 7.35 | 10 | 4.21 | 0.21 | 8.22 | 4.01 |
| 29 | NASO | 10 | 16.79 | -45.71 | 79.30 | 62.50 | 10 | 17.93 | -22.98 | 58.85 | 40.91 | 10 | 13.29 | -43.36 | 69.93 | 56.64 | 10 | 13.67 | -26.00 | 53.35 | 39.67 |
| 30 | ORO | 10 | -8.35 | -68.43 | 51.73 | 60.08 | 10 | -69.47 | -175.44 | 36.50 | 105.97 | 10 | -34.70 | -119.64 | 50.25 | 84.95 | 10 | -47.18 | -108.70 | 14.33 | 61.51 |
| 31 | HC | 10 | 1.65 | -15.09 | 18.39 | 16.74 | 10 | -6.15 | -22.44 | 10.14 | 16.29 | 10 | 2.67 | -17.82 | 23.17 | 20.50 | 10 | 2.94 | -15.94 | 21.82 | 18.88 |

| Relative Difference | | Observer 0* | | | | | Observer 2 | | | | | Observer 3 | | | | | Automated | | | | |
|---|---|---|---|---|---|---|---|---|---|---|---|---|---|---|---|---|---|---|---|---|---|
| ID | Variable | N_Obs | Mean | Lower_CI | Upper_CI | rel LoA range (+/- mean) | N_Obs | Mean | Lower_CI | Upper_CI | rel LoA range (+/- mean) | N_Obs | Mean | Lower_CI | Upper_CI | rel LoA range (+/- mean) | N_Obs | Mean | Lower_CI | Upper_CI | rel LoA range (+/- mean) |
| 1 | ASBL | 10 | 5.20 | -1.94 | 12.34 | 7.14 | 10 | 0.32 | -5.43 | 6.07 | 5.75 | 10 | 0.24 | -4.86 | 5.34 | 5.10 | 10 | 2.13 | -3.46 | 7.72 | 5.59 |
| 2 | PSBL | 10 | 0.36 | -9.87 | 10.58 | 10.22 | 10 | -4.16 | -18.69 | 10.38 | 14.54 | 10 | -0.19 | -13.61 | 13.23 | 13.42 | 10 | -1.36 | -11.63 | 8.91 | 10.27 |
| 3 | HPL | 10 | -4.30 | -24.46 | 15.87 | 20.16 | 10 | -6.58 | -42.75 | 29.60 | 36.18 | 10 | -13.91 | -48.32 | 20.50 | 34.41 | 10 | -2.70 | -17.71 | 12.31 | 15.01 |
| 4 | VPL | 10 | -0.21 | -13.16 | 12.75 | 12.95 | 10 | 1.76 | -22.87 | 26.39 | 24.63 | 10 | -4.71 | -21.98 | 12.56 | 17.27 | 10 | -5.58 | -13.48 | 2.32 | 7.90 |
| 5 | PAW | 10 | -2.75 | -19.79 | 14.28 | 17.04 | 10 | -8.69 | -24.70 | 7.32 | 16.01 | 10 | -11.18 | -25.98 | 3.62 | 14.80 | 10 | -1.26 | -18.81 | 16.29 | 17.55 |
| 6 | Cho_r | 6 | 9.50 | -4.04 | 23.03 | 13.53 | 7 | -2.76 | -30.54 | 25.02 | 27.78 | 6 | 19.99 | -32.33 | 72.31 | 52.32 | 7 | 2.18 | -15.43 | 19.78 | 17.61 |
| 7 | Cho_l | 6 | 0.27 | -17.90 | 18.43 | 18.16 | 7 | -3.74 | -29.71 | 22.22 | 25.96 | 6 | 16.30 | -31.80 | 64.39 | 48.10 | 7 | -4.47 | -27.69 | 18.74 | 23.22 |
| 8 | NPW | 6 | 2.21 | -30.95 | 35.38 | 33.17 | 7 | 0.86 | -23.75 | 25.46 | 24.60 | 6 | 17.82 | -34.58 | 70.22 | 52.40 | 7 | 14.20 | -4.58 | 32.97 | 18.78 |
| 9 | NB | 7 | 8.21 | -51.13 | 67.54 | 59.34 | 8 | 29.36 | -6.32 | 65.04 | 35.68 | 9 | -0.35 | -50.25 | 49.56 | 49.90 | 9 | 19.69 | -0.06 | 39.44 | 19.75 |
| 10 | PNTh | 10 | -0.69 | -67.52 | 66.13 | 66.83 | 10 | -18.60 | -138.39 | 101.19 | 119.79 | 10 | -12.10 | -90.88 | 66.68 | 78.78 | 10 | -2.88 | -59.90 | 54.14 | 57.02 |
| 11 | OFD | 10 | 0.24 | -4.60 | 5.07 | 4.84 | 10 | -1.16 | -6.58 | 4.26 | 5.42 | 10 | 0.32 | -6.65 | 7.29 | 6.97 | 10 | 0.36 | -4.77 | 5.49 | 5.13 |
| 12 | BPD | 10 | 1.48 | -7.16 | 10.11 | 8.64 | 10 | -0.93 | -8.90 | 7.04 | 7.97 | 10 | -0.73 | -8.98 | 7.52 | 8.25 | 10 | 2.24 | -5.04 | 9.52 | 7.28 |
| 13 | MCh | 10 | 2.23 | -4.62 | 9.09 | 6.86 | 10 | 1.43 | -4.32 | 7.18 | 5.75 | 10 | 0.99 | -6.24 | 8.21 | 7.23 | 10 | -0.47 | -7.36 | 6.42 | 6.89 |
| 14 | OD_R | 10 | -4.12 | -17.23 | 8.98 | 13.11 | 10 | -5.34 | -15.77 | 5.09 | 10.43 | 10 | -5.82 | -19.01 | 7.37 | 13.19 | 10 | 1.01 | -9.43 | 11.46 | 10.45 |
| 15 | OD_L | 10 | -2.76 | -10.67 | 5.15 | 7.91 | 10 | -3.90 | -16.01 | 8.20 | 12.11 | 10 | -6.30 | -14.83 | 2.23 | 8.53 | 10 | 0.56 | -5.78 | 6.90 | 6.34 |
| 16 | IOD | 10 | 2.52 | -11.81 | 16.84 | 14.32 | 10 | -7.90 | -27.23 | 11.42 | 19.33 | 10 | 3.56 | -11.96 | 19.07 | 15.51 | 10 | 0.46 | -13.08 | 13.99 | 13.54 |
| 17 | BOD | 10 | -0.65 | -6.53 | 5.22 | 5.88 | 10 | -1.38 | -7.28 | 4.53 | 5.91 | 10 | -2.05 | -8.24 | 4.13 | 6.18 | 10 | 0.98 | -4.02 | 5.99 | 5.01 |
| 18 | MxW | 10 | 4.81 | -3.29 | 12.91 | 8.10 | 10 | 2.59 | -12.26 | 17.45 | 14.86 | 10 | -3.82 | -23.17 | 15.52 | 19.34 | 10 | 7.67 | -0.42 | 15.76 | 8.09 |
| 19 | MdW | 10 | 4.55 | -5.27 | 14.37 | 9.82 | 10 | 0.93 | -3.14 | 4.99 | 4.06 | 10 | -2.63 | -9.97 | 4.70 | 7.34 | 10 | 14.62 | 5.02 | 24.22 | 9.60 |
| 20 | CBA1 | 10 | 0.41 | -3.55 | 4.38 | 3.96 | 10 | 1.61 | -4.20 | 7.42 | 5.81 | 10 | -0.08 | -5.78 | 5.63 | 5.71 | 10 | 0.27 | -5.37 | 5.91 | 5.64 |
| 21 | CBA2 | 10 | -7.61 | -46.82 | 31.61 | 39.21 | 10 | -2.64 | -40.13 | 34.85 | 37.49 | 10 | 6.36 | -17.86 | 30.57 | 24.22 | 10 | -9.89 | -30.61 | 10.84 | 20.72 |
| 22 | FMA | 10 | 0.43 | -11.48 | 12.34 | 11.91 | 10 | 2.25 | -8.03 | 12.53 | 10.28 | 10 | -5.24 | -20.49 | 10.01 | 15.25 | 10 | 0.06 | -11.74 | 11.87 | 11.81 |
| 23 | MNMA | 10 | -108.17 | -266.13 | 49.79 | 157.96 | 10 | -41.68 | -172.55 | 89.20 | 130.88 | 10 | -106.79 | -275.52 | 61.94 | 168.73 | 10 | -81.55 | -224.31 | 61.21 | 142.76 |
| 24 | IFA | 10 | 1.98 | -23.19 | 27.16 | 25.17 | 10 | 7.69 | -20.03 | 35.41 | 27.72 | 10 | -4.95 | -42.06 | 32.17 | 37.11 | 10 | 4.38 | -18.75 | 27.52 | 23.13 |
| 25 | PAH | 10 | -52.80 | -163.65 | 58.05 | 110.85 | 10 | -56.26 | -175.35 | 62.83 | 119.09 | 10 | -47.42 | -224.12 | 129.27 | 176.69 | 10 | -18.30 | -144.68 | 108.09 | 126.38 |
| 26 | ChoH | 6 | 33.23 | -7.10 | 73.56 | 40.33 | 6 | 20.17 | -35.33 | 75.68 | 55.50 | 6 | -15.54 | -71.05 | 39.97 | 55.51 | 6 | 10.91 | -62.52 | 84.35 | 73.43 |
| 27 | MXL | 10 | -4.69 | -24.68 | 15.29 | 19.99 | 10 | 1.07 | -13.58 | 15.72 | 14.65 | 10 | 9.50 | -35.23 | 54.24 | 44.73 | 10 | -3.46 | -16.83 | 9.90 | 13.36 |
| 28 | MDL | 10 | 11.62 | -10.17 | 33.41 | 21.79 | 10 | 0.83 | -14.45 | 16.11 | 15.28 | 10 | 3.31 | -27.34 | 33.97 | 30.66 | 10 | 17.31 | 4.32 | 30.30 | 12.99 |
| 29 | NASO | 10 | 20.20 | -42.15 | 82.55 | 62.35 | 10 | 17.76 | -19.50 | 55.03 | 37.26 | 10 | 10.62 | -46.51 | 67.75 | 57.13 | 10 | 12.14 | -33.73 | 58.01 | 45.87 |
| 30 | ORO | 10 | -10.05 | -73.24 | 53.14 | 63.19 | 10 | -62.32 | -213.87 | 89.23 | 151.55 | 10 | -28.78 | -107.80 | 50.24 | 79.02 | 10 | -39.77 | -110.85 | 31.32 | 71.08 |
| 31 | HC | 10 | 0.55 | -5.10 | 6.21 | 5.65 | 10 | -2.05 | -7.49 | 3.39 | 5.44 | 10 | 0.93 | -5.86 | 7.72 | 6.79 | 10 | 0.98 | -5.29 | 7.25 | 6.27 |

Mean within +/- 10% or LoA within +/- 20%

Automated variation higher human observers

**Figure 10.** Results of absolute (upper table) and relative (lower table) differences with observer 0 (manual landmark based indirect measurement), observer 2 and 3 (direct manual measurement) and automated results compared to a single expert radiologist (direct manual measurement. Green represents relative mean difference or <10% or relative limits of agreement (+/− mean) of <20%. Orange represents automated measurements with more variation than any one manual observer.

*average measures

| | Intraobserver | | | | | Interobserver | | | | | Intermethod | | | | | ICC threshold criteria: <0.50 = poor; 0.50 -0.75 = moderate; 0.75 - 0.90 = good; >0.90 = excellent (Koo and Li, 2016) | | |
|---|---|---|---|---|---|---|---|---|---|---|---|---|---|---|---|---|---|---|
| Variable | n | Cronbach's Alpha | Intraclass Correlation* | 95% Confidence Lower Bound | Upper Bound | n | Cronbach's Alpha | Intraclass Correlation* | 95% Confidence Lower Bound | Upper Bound | n | Cronbach's Alpha | Intraclass Correlation* | 95% Confidence Lower Bound | Upper Bound | Intrasobserver (indirect measurement | Interobserver (direct measurement | Intermethod (direct, indirect, automated |
| ASBL | 10 | 0.952 | **0.952** | 0.818 | 0.988 | 10 | 0.936 | **0.942** | 0.829 | 0.984 | 10 | 0.963 | **0.926** | 0.770 | 0.980 | Excellent | Excellent | Excellent |
| PSBL | 10 | 0.747 | **0.754** | 0.033 | 0.938 | 10 | 0.889 | **0.879** | 0.659 | 0.967 | 10 | 0.918 | **0.915** | 0.793 | 0.976 | Good | Good | Excellent |
| HPL | 10 | 0.795 | **0.811** | 0.190 | 0.954 | 10 | 0.434 | **0.392** | -0.463 | 0.822 | 10 | 0.640 | **0.603** | 0.099 | 0.882 | Good | n/a | n/a |
| VPL | 10 | 0.529 | **0.431** | -0.476 | 0.836 | 10 | 0.418 | **0.397** | -0.569 | 0.829 | 10 | 0.588 | **0.554** | -0.009 | 0.867 | n/a | n/a | n/a |
| PAW | 10 | 0.973 | **0.975** | 0.901 | 0.994 | 10 | 0.899 | **0.791** | 0.256 | 0.947 | 10 | 0.926 | **0.856** | 0.596 | 0.960 | Excellent | Good | Good |
| Cho_r | 5 | 0.741 | **0.769** | -1.788 | 0.977 | 6 | 0.431 | **0.275** | -0.386 | 0.840 | 5 | 0.834 | **0.660** | 0.130 | 0.953 | Good | n/a | Moderate |
| Cho_l | 5 | 0.750 | **0.429** | -0.257 | 0.909 | 6 | 0.627 | **0.396** | -0.206 | 0.870 | 5 | 0.712 | **0.464** | -0.054 | 0.911 | Good | n/a | Poor |
| NPW | 6 | 0.959 | **0.964** | 0.759 | 0.995 | 6 | -0.104 | **-0.029** | -0.326 | 0.608 | 5 | -1.419 | **-0.292** | -0.616 | 0.566 | Excellent | n/a | n/a |
| NB | 7 | 0.417 | **0.429** | -2.607 | 0.903 | 8 | -0.578 | **-0.254** | -0.964 | 0.553 | 5 | 0.458 | **0.448** | -0.864 | 0.934 | n/a | n/a | n/a |
| PNTh | 10 | 0.560 | **0.521** | -0.522 | 0.873 | 10 | 0.345 | **0.361** | -1.043 | 0.833 | 10 | 0.507 | **0.514** | -0.223 | 0.863 | n/a | n/a | n/a |
| OFD | 10 | 0.981 | **0.981** | 0.927 | 0.995 | 10 | 0.974 | **0.974** | 0.926 | 0.993 | 10 | 0.982 | **0.982** | 0.956 | 0.995 | Excellent | Excellent | Excellent |
| BPD | 10 | 0.994 | **0.987** | 0.765 | 0.998 | 10 | 0.933 | **0.937** | 0.818 | 0.983 | 10 | 0.923 | **0.916** | 0.795 | 0.976 | Excellent | Excellent | Excellent |
| MCh | 10 | 0.959 | **0.927** | 0.446 | 0.984 | 10 | 0.958 | **0.958** | 0.882 | 0.989 | 10 | 0.971 | **0.968** | 0.921 | 0.991 | Excellent | Excellent | Excellent |
| OD_R | 10 | 0.943 | **0.926** | 0.661 | 0.982 | 10 | 0.773 | **0.691** | 0.180 | 0.912 | 10 | 0.895 | **0.837** | 0.584 | 0.953 | Excellent | Moderate | Good |
| OD_L | 10 | 0.866 | **0.774** | -0.058 | 0.947 | 10 | 0.846 | **0.793** | 0.403 | 0.943 | 10 | 0.916 | **0.878** | 0.687 | 0.966 | Good | Good | Good |
| IOD | 10 | 0.909 | **0.861** | 0.284 | 0.968 | 10 | 0.685 | **0.556** | -0.033 | 0.866 | 10 | 0.824 | **0.750** | 0.413 | 0.926 | Good | n/a | Moderate |
| BOD | 10 | 0.955 | **0.943** | 0.743 | 0.986 | 10 | 0.941 | **0.930** | 0.793 | 0.981 | 10 | 0.960 | **0.950** | 0.873 | 0.986 | Excellent | Excellent | Excellent |
| MxW | 10 | 0.936 | **0.942** | 0.762 | 0.986 | 10 | 0.428 | **0.411** | -0.559 | 0.835 | 10 | 0.796 | **0.727** | 0.372 | 0.919 | Excellent | n/a | Moderate |
| MdW | 10 | 0.823 | **0.833** | 0.323 | 0.959 | 10 | 0.934 | **0.904** | 0.677 | 0.975 | 10 | 0.901 | **0.635** | 0.176 | 0.888 | Good | Good | Moderate |
| CBA1 | 10 | 0.555 | **0.581** | -0.965 | 0.899 | 10 | 0.915 | **0.907** | 0.735 | 0.974 | 10 | 0.934 | **0.934** | 0.839 | 0.981 | n/a | Excellent | Excellent |
| CBA2 | 10 | 0.855 | **0.745** | -0.139 | 0.822 | 10 | -0.332 | **-0.300** | -2.319 | 0.627 | 10 | 0.154 | **0.130** | -0.755 | 0.722 | Moderate | n/a | n/a |
| FMA | 10 | 0.607 | **0.602** | -0.474 | 0.940 | 10 | 0.777 | **0.692** | 0.181 | 0.913 | 10 | 0.817 | **0.777** | 0.475 | 0.935 | n/a | Moderate | Good |
| MNMA | 10 | 0.612 | **0.619** | -0.514 | 0.905 | 10 | -0.557 | **-0.176** | -0.581 | 0.423 | 10 | 0.197 | **0.090** | -0.246 | 0.574 | n/a | n/a | n/a |
| IFA | 10 | 0.863 | **0.872** | 0.479 | 0.968 | 10 | 0.730 | **0.701** | 0.201 | 0.916 | 10 | 0.760 | **0.747** | 0.395 | 0.927 | Good | Moderate | Good |
| PAH | 10 | 0.890 | **0.827** | 0.138 | 0.960 | 10 | 0.082 | **0.067** | -1.011 | 0.709 | 10 | 0.044 | **0.036** | -0.880 | 0.685 | Good | n/a | n/a |
| ChoH | 10 | 0.440 | **0.445** | -1.226 | 0.862 | 6 | 0.465 | **0.375** | -0.565 | 0.886 | 6 | 0.617 | **0.505** | -0.139 | 0.907 | n/a | n/a | n/a |
| MXL | 10 | 0.555 | **0.556** | -0.704 | 0.883 | 10 | 0.543 | **0.528** | -0.274 | 0.869 | 10 | 0.501 | **0.471** | -0.195 | 0.843 | n/a | n/a | n/a |
| MDL | 10 | 0.654 | **0.647** | -0.299 | 0.910 | 10 | 0.442 | **0.448** | -0.634 | 0.852 | 10 | 0.674 | **0.515** | 0.057 | 0.839 | n/a | n/a | n/a |
| NASO | 10 | 0.847 | **0.833** | 0.384 | 0.958 | 10 | -0.219 | **-0.168** | -1.421 | 0.625 | 10 | 0.316 | **0.309** | -0.647 | 0.800 | Good | n/a | n/a |
| ORO | 10 | 0.671 | **0.675** | -0.265 | 0.919 | 10 | 0.601 | **0.431** | -0.146 | 0.813 | 10 | 0.745 | **0.618** | 0.187 | 0.880 | n/a | n/a | Moderate |
| HC | 10 | 0.988 | **0.979** | 0.839 | 0.996 | 10 | 0.968 | **0.953** | 0.831 | 0.988 | 10 | 0.964 | **0.959** | 0.899 | 0.988 | Excellent | Excellent | Excellent |

**Figure 11.** Table of Intraclass Correlation Coefficient (ICC) results. Green is and excellent or good agreement, yellow a moderate agreement, red is poor agreement (n.b. ICC threshold criteria interpreted only if Cronbach's Alpha is >0.70).

**(a)** Confidence in 3D landmark placement (n=35) between first and second repeated observation

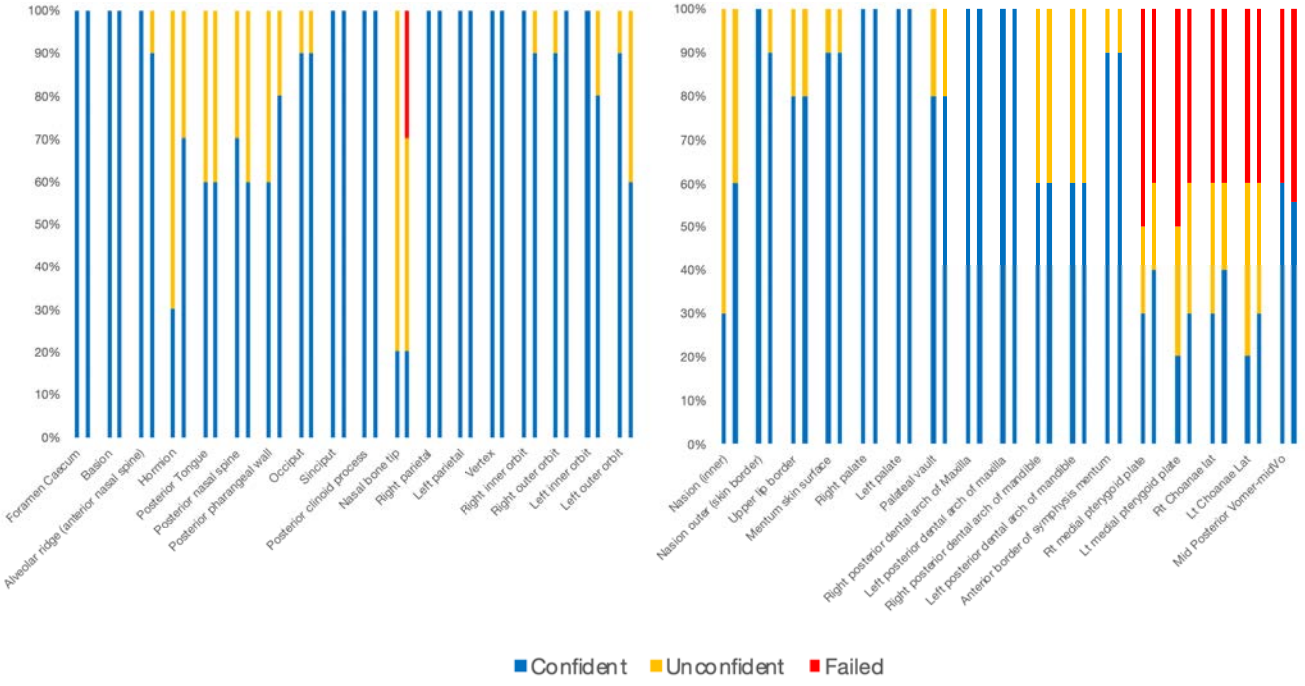


**(b)** Boxplots of average confidence for 3 raters for obtaining measurements (scale: 0 = failed, 1 = unconfident, 2 = Confident). * Represents case number outliers, see S2 table 4. Note, small boxes at point 2 on the y-axis represents no variation.

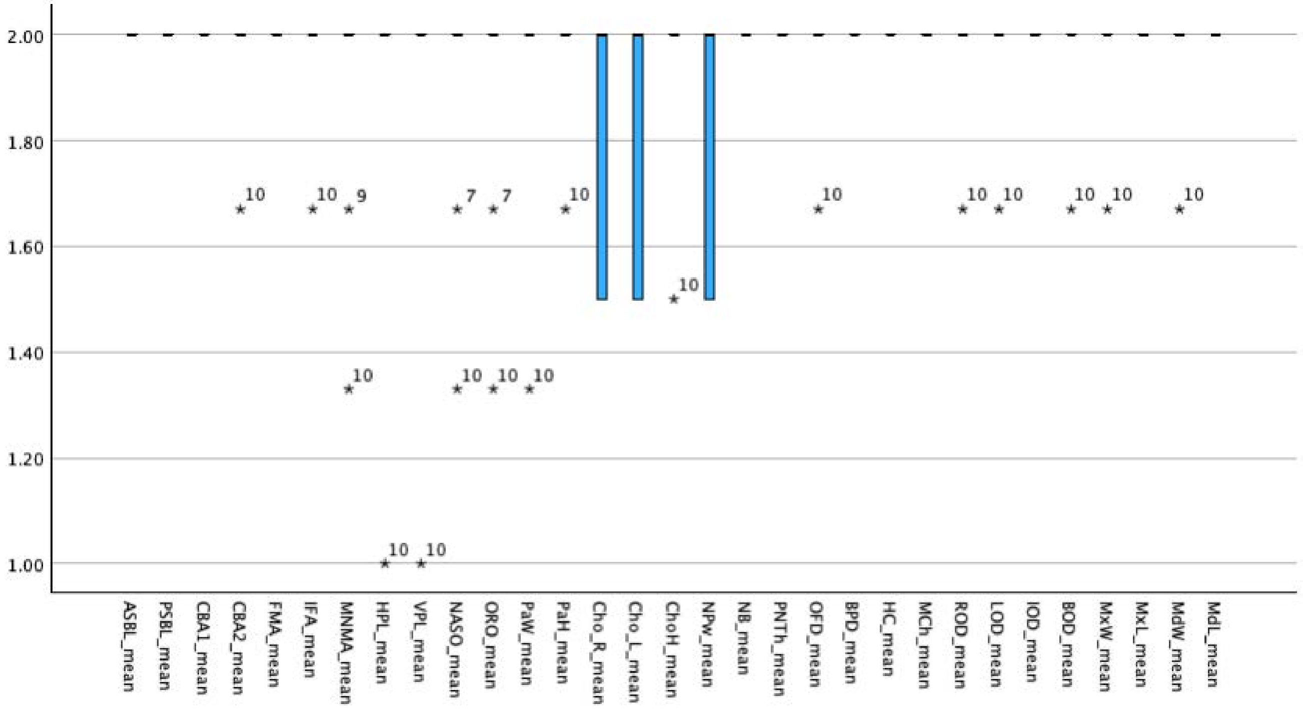


**Figure 12.** Human Observer Diagnostic Confidence of Landmark placement and measurement accuracy. a. Observer 0 ( repeated measures). b. Average of observers 1 to 3 (each performing single independent measurements

**Figure 13.** GA-matched fetal MRI datasets used in the control (104) and T21 (24) comparison a. GA distribution b. MRI protocols and fetal sex

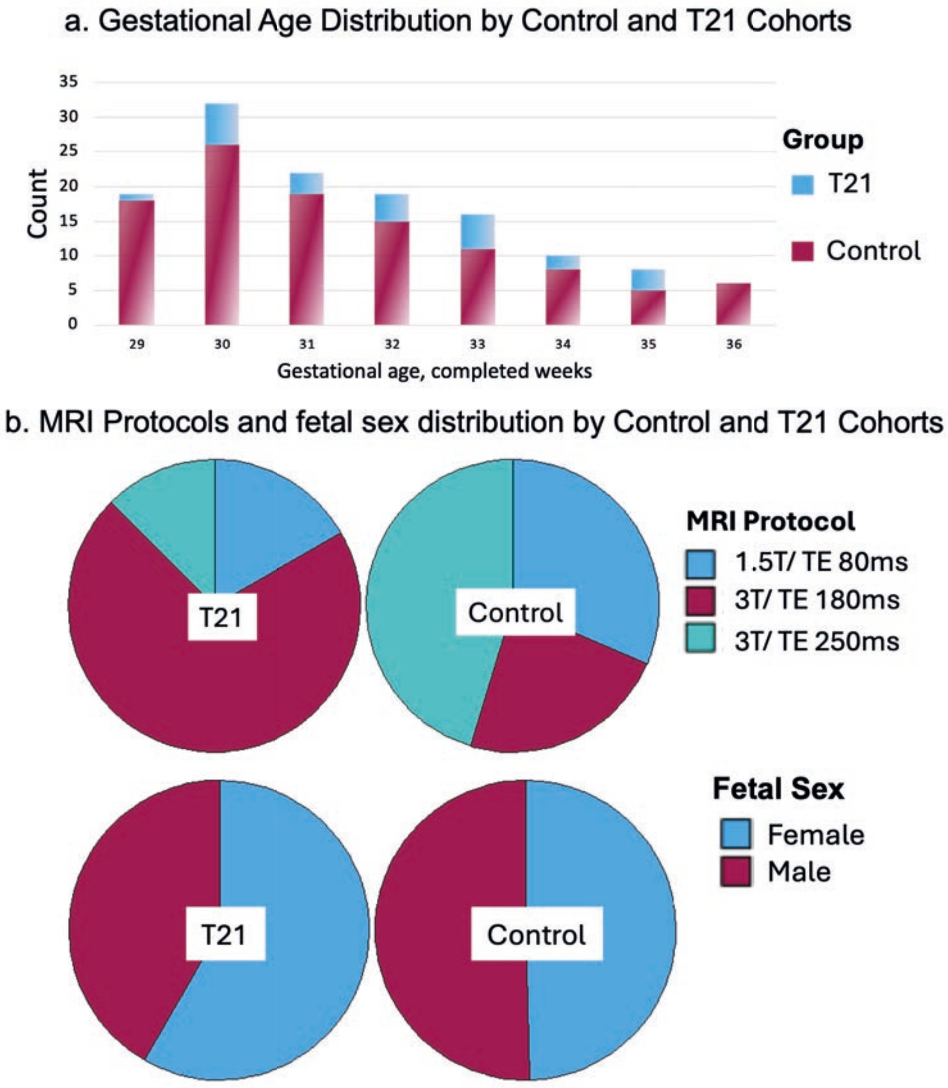

**Figure 14.** Chart demostrating the proportion of biometrics that fell outside of normative range (5th - 95th percentile) for each T21 subject, upper chart: for most significant biometrics and lower chart all biometrics.

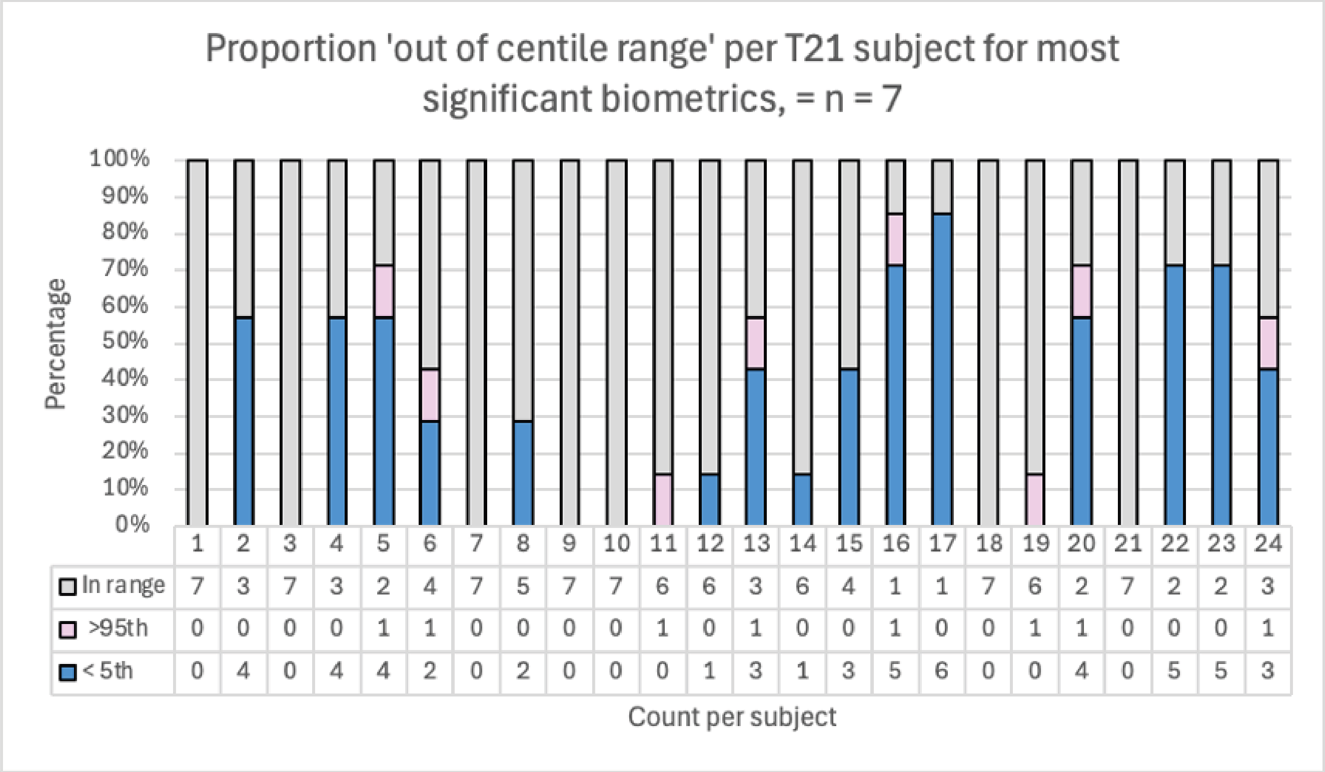


| | 1 | 2 | 3 | 4 | 5 | 6 | 7 | 8 | 9 | 10 | 11 | 12 | 13 | 14 | 15 | 16 | 17 | 18 | 19 | 20 | 21 | 22 | 23 | 24 |
|---|---|---|---|---|---|---|---|---|---|---|---|---|---|---|---|---|---|---|---|---|---|---|---|---|
| In range | 7 | 3 | 7 | 3 | 2 | 4 | 7 | 5 | 7 | 7 | 6 | 6 | 3 | 6 | 4 | 1 | 1 | 7 | 6 | 2 | 7 | 2 | 2 | 3 |
| >95th | 0 | 0 | 0 | 0 | 1 | 1 | 0 | 0 | 0 | 0 | 1 | 0 | 1 | 0 | 0 | 1 | 0 | 0 | 1 | 1 | 0 | 0 | 0 | 1 |
| < 5th | 0 | 4 | 0 | 4 | 4 | 2 | 0 | 2 | 0 | 0 | 0 | 1 | 3 | 1 | 3 | 5 | 6 | 0 | 0 | 4 | 0 | 5 | 5 | 3 |

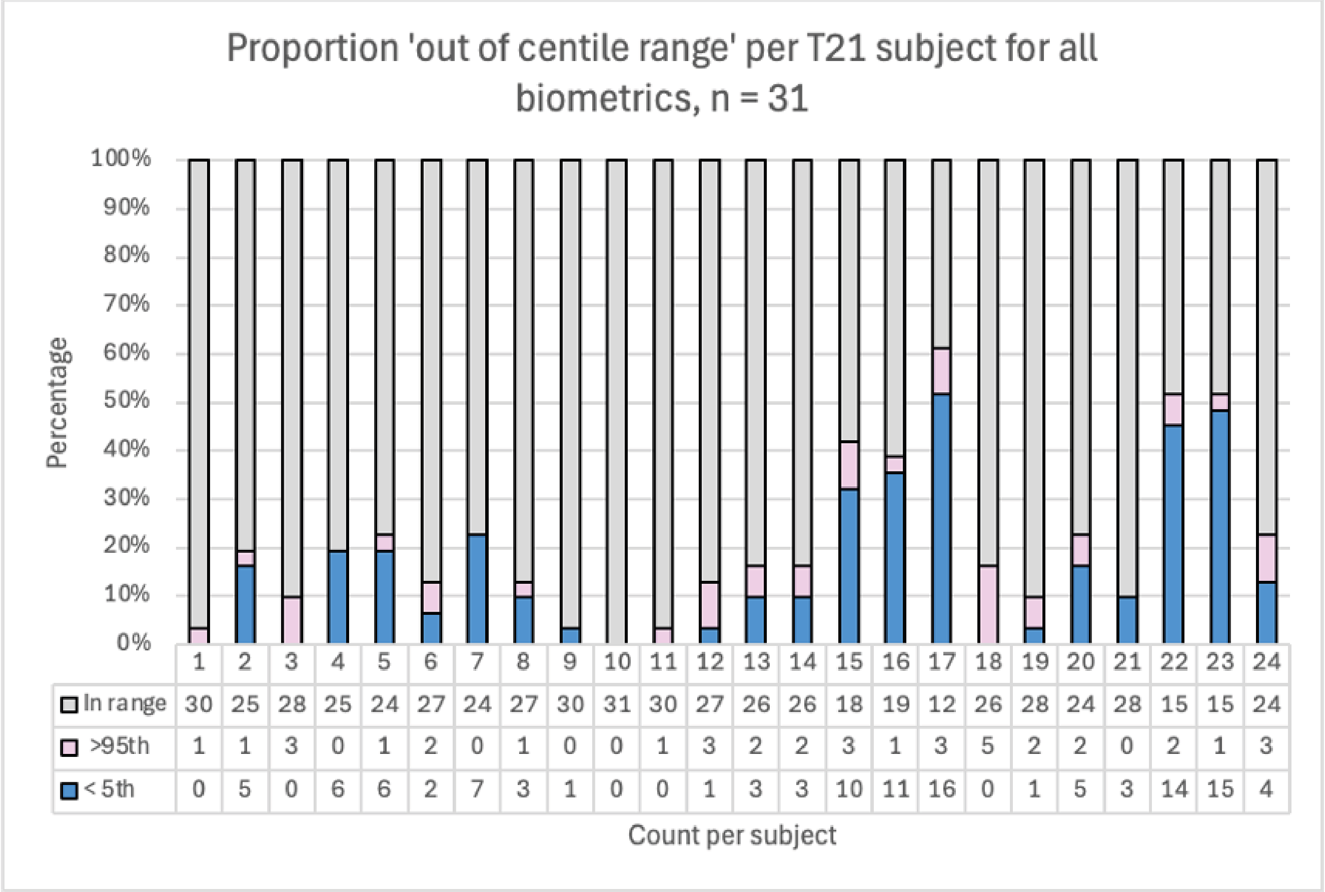


| | 1 | 2 | 3 | 4 | 5 | 6 | 7 | 8 | 9 | 10 | 11 | 12 | 13 | 14 | 15 | 16 | 17 | 18 | 19 | 20 | 21 | 22 | 23 | 24 |
|---|---|---|---|---|---|---|---|---|---|---|---|---|---|---|---|---|---|---|---|---|---|---|---|---|
| In range | 30 | 25 | 28 | 25 | 24 | 27 | 24 | 27 | 30 | 31 | 30 | 27 | 26 | 26 | 18 | 19 | 12 | 26 | 28 | 24 | 28 | 15 | 15 | 24 |
| >95th | 1 | 1 | 3 | 0 | 1 | 2 | 0 | 1 | 0 | 0 | 1 | 3 | 2 | 2 | 3 | 1 | 3 | 5 | 2 | 2 | 0 | 2 | 1 | 3 |
| < 5th | 0 | 5 | 0 | 6 | 6 | 2 | 7 | 3 | 1 | 0 | 0 | 1 | 3 | 3 | 10 | 11 | 16 | 0 | 1 | 5 | 3 | 14 | 15 | 4 |
