## Supplementary Tables S2 for "Automated Craniofacial Biometry with 3D T2w Fetal MRI"

### S2: Supplementary Tables

**Table 2.** 3D Anatomical Landmarks (*Landmark not used for biometry extraction)

| Label number | Anatomical point | Abbr. | Description |
|---|---|---|---|
| 1 | Foramen Caecum | Fc | The midline point marking the pit between the fetal crista galli and the endocranial wall of the frontal bone |
| 2 | Basion | Ba | The midline point on the anterior margin of the foramen magnum |
| 3 | Alveolar ridge (anterior nasal spine) | ANS | The midline central point on the tip of the alveolar ridge (also synonymous with anterior nasal spine, ANS) |
| 4 | Hormion | H | The posterior-most midline point on the junction between the ventral surface of the sphenoid and the vomeral root (also, point located at the intersection between the perpendicular line to S-Ba from PNS and the cranial base) |
| 5 | Posterior Tongue | TP | Posterior most aspect of the tongue in the midline |
| 6 | Posterior nasal spine | PNS | The midline point of the superior surface of the hard palate where the horizontal aspect of the palatine bone meets the posterior most aspect of the vomer bone. |
| 7 | Posterior pharangeal wall | PPW | The posterior point of the pharanyx on a linear line contiguous with the hard palate length (ANS-PNS) |
| 8 | Occiput | Oc | Posterior most skull border in axial plane at level of thalami |
| 9* | Cervical axis | C1 | The midline point on the first vertebral body anterior to the basion point |
| 10 | Sinciput | Si | Anterior most skull border in axial plane at level of thalami |
| 11 | Posterior clinoid process | PCP | The midline point on the posteior raised tuberculum sella of the body of the sphenoid |
| 12 | Nasal bone tip | NB | Inferior most tip of the nasal bone |
| 13* | Nose tip | NBT | Tip of the superficial surface of nasal soft tissue |
| 14* | Tongue root | TR | Most inferior posterior muscular part of the tongue (estimated site of hyoid bone) |
| 15 | Right parietal | Pt_r | Right lateral most skull border in axial plane at level of SI-Oc plane (OFD/BPD) |
| 16 | Left parietal | Pt_l | Left lateral most skull border in axial plane at level of Si-Oc plane (OFD/BPD) |
| 17 | Vertex | Ve | Most superior skull border in a midline sagittal plane (at level of cerebral peduncles when oriented to brain orthgonal planes in coronal) |
| 18 | Right palate | pa_r | Right mid-inferior border of posterior most maxillary molar in coronal plane (when oriented for coronal facial views) |

Continued on next page

Table 2 – Continued from previous page

| Label number | Anatomical point | Abbr. | Description |
|---|---|---|---|
| 19 | Left palate | pa_l | Left mid-inferior border of posterior most maxillary molar in coronal plane (when oriented for coronal facial views) |
| 20 | Palateal vault | PaV | Midline point in coronal plane at level of posterior most maxillary molar at the inferior border of the hard palate |
| 21 | Right posterior dental arch of Maxilla | Mx_r | Posterior point of last right molar tooth socket at the level of the widest point of the maxillary dental arch |
| 22 | Left posterior dental arch of maxilla | Mx_l | Posterior point of last left molar tooth socket at the level of the widest point of the maxillary dental arch |
| 23 | Right posterior dental arch of mandible | man_r | Posterior point of last right molar tooth socket at the level of the widest point of the mandibular dental arch (at the level of the masseter on its entrance point to the mandible) |
| 24 | Left posterior dental arch of mandible | man_l | Posterior point of last left molar tooth socket at the level of the widest point of the mandibular dental arch (at the level of the masseter on its entrance point to the mandible) |
| 25 | Anterior border of symphysis mentum | Me | Midline point at the level of the symphysis mentis of the mandible |
| 26* | Right Superior external ear | hel_r | Right most superior point of the helix (external ear) |
| 27* | Right inferior external ear | lob_r | Right most interior point of the auricular lobule (external ear) |
| 28* | Right external auditory meatus | EAM_r | Midpoint of the right external auditory meatus (between the tragus and the concha) |
| 29* | Left Superior external ear | hel_l | Left most superior point of the helix (external ear) |
| 30* | Left inferior external ear | lob-l | Left most interior point of the auricular lobule (external ear) |
| 31* | Left external auditory meatus | EAM_l | Midpoint of the left external auditory meatus (between the tragus and the concha) |
| 32 | Right inner orbit | RO_in | Right inner border of the orbital globe (i.e. medial outer edge of sclera) at widest point in axial view |
| 33 | Right outer orbit | RO_o | Right outer border of the orbital globe (i.e. lateral outer edge of sclera) at widest point in axial view |
| 34 | Left inner orbit | LO_in | Left inner border of the orbital globe (i.e. medial outer edge of sclera) at widest point in axial view |
| 35 | Left outer orbit | LO_o | Left outer border of the orbital globe (i.e. lateral outer edge of sclera) at widest point in axial view |
| 36 | Nasion (inner) | NaIn | Inner border of nasion |
| 37 | Nasion outer (skin border) | NaO | Outer skin border of nasion |
| 38* | Nuchal fold inner | NF_in | Nuchal inner border (medial subcutaneous layer) |
| 39* | Nuchal fold outer (skin border) | NF_o | Nuchal outer border (posterior border subcutaneous layer) |
| 40 | Upper lip border | Lip | Skin border of the upper lip midline |

Continued on next page

Table 2 – Continued from previous page

| Label number | Anatomical point | Abbr. | Description |
|---|---|---|---|
| 41 | Mentum skin surface | Chin | Skin border of the chin midline |
| 42* | opisthion | Op | Midline posterior border of the foramen magnum |
| 43* | Right Lateral Foramen Magnum | FM_r | Right lateral border of the foramen magnum |
| 44* | Left lateral Foramen Magnum | FM_l | Left lateral inner border of the foramen Magnum |
| 45* | Tongue tip | TT | Most anterior part of tongue in the midline |
| 46 | Rt medial pterygoid plate | MPP_r | Point on the right posterior lateral border of the nasal labrinth at a level of the vomer |
| 47 | Lt medial pterygoid plate | MPP_l | Point on the left posterior lateral border of the nasal labrinth at a level of the vomer |
| 48 | Rt Choanae lat | Cho_r | Right posterior lateral border of the anterior nasopharyneal space, posterior to the vomer |
| 49 | Lt Choanae Lat | Cho_l | Left posterior lateral border of the anterior nasopharyneal space, posterior to the vomer |
| 50 | Mid Posterior Vomer-midVo | Vo | Point in the midline of the vomer level with the right and left medial pterygoid plate |

**Table 3.** Formalised measurement definitions for landmark-based craniofacial biometry protocol with 3D T2w fetal MRI.

| Anatomical group | Measure Number | Measurement name | Abbreviation | Landmark points used |
|---|---|---|---|---|
| | 1 | Anterior skull base length | ASBL | Fc (1), PCP (11) |
| 1. Base of skull | 2 | Posterior skull base length | PSBL | PCP (11), Ba (2) |
| | 3 | Internal cranial Base angle (°) | CBA1 | Fc(1), PCP (11), Ba (2) |
| | 4 | External cranial base angle (°) | CBA2 | PNS (6), H (4), Ba (2) |
| 2. Facial angles | 5 | Fronto maxillary angle | FMA | Si (10), Fc (1), PCP (11) |
| | 6 | Inferior facial angle | IFA | PCP (11), Fc (1), Lip (40), Chin (41) |
| | 7 | Maxillary nasion mandibular angle | MNMA | ANS (3), Fc (1), Me (25) |
| 3. Oropharangeal | 8 | Hard palate length | HPL | ANS (3), PNS (6) |
| | 9 | Velopharangeal length | VPL | ANS (3), PNS (6), PPW (7) |
| | 10 | Nasopharyngeal area (mm2) | NASO | PNS (6), H (4), Ba (2) |
| | 11 | Oropharyngeal area (mm2) | ORO | PNS (6), Ba (2), TP (5) |
| | 12 | Palatal width | PaW | Pa (Rt/Lt) (18,19) |
| | 13 | Palatal height | PaH | Pav (20), RtPa, Lt Pa (18, 19) |
| 4. Nasal | 14-15 | Choanae width (rt/Lt) | Cho_R/Cho_L | MPP (Rt/Lt) (46, 47), Vo (50) |
| | 16 | Choanae height (mm) | ChoH | H (4), ANS (3), PNS (6) |
| | 17 | Nasopharynx width | NPw | Cho (Lt/Rt) (48, 49) |
| | 18 | Nasal bone | NB | NaIn (36), NB (12) |
| | 19 | Prenasal thickness | PNTh | NaIn (36), NaO (37) |
| | 20 | Occipital frontal diameter | OFD | Si (10), Oc (8) |
| 5. Cranial vault | 21 | Bi parietal diameter | BPD | Pt (Rt/Lt) (15/16) |
| | 22 | Head circumference | HC | SI (10), Oc (8), RtPt (15), LtPt (16) |
| | 23 | Maximum cranial height | MCh | Ve (17), Ba (2) |
| | 24-25 | Orbital distance | ROD/LOD | RO/LO_in (33/35), RO/LO_o (34/36) |
| 6. Orbits | 26 | Interoccular distance | IOD | RO_in (33), LO_i (35) |
| | 27 | Biocular distance | BOD | RO/LO_o (34/36) |
| 7. Maxilla | 28 | Maxillary width | MxW | RtMx (21), LtMx (22) |
| | 29 | Maxillary length | MxL | ANS (3), RtMx (21), LtMx (22) |
| 8. Mandible | 30 | Mandibular Width | MdW | RtMd (23), LtMd (24) |
| | 31 | Mandibular length | MdL | Me (25), RtMd (23), LtMd (24) |

**Table 4.** Demographics of the 10 cases selected for quantitative evaluation of the proposed biometry protocol and pipeline.

| ID | GA | Group | Sex | TE | Field Strength | HeadSVR quality |
|---|---|---|---|---|---|---|
| 1 | 29.86 | T21 | Male | 180ms | 3.0T | Good |
| 2 | 35.43 | T21 | Male | 180ms | 3.0T | Moderate |
| 3 | 32.71 | T21 | Male | 250ms | 3.0T | Moderate |
| 4 | 33.71 | T21 | Female | 80ms | 1.5T | Good |
| 5 | 32.71 | T21 | Female | 80ms | 1.5T | Excellent |
| 6 | 31.86 | Control | Male | 80ms | 1.5T | Moderate |
| 7 | 32.86 | Control | Female | 80ms | 1.5T | Good |
| 8 | 29.43 | Control | Female | 80ms | 1.5T | Moderate |
| 9 | 30.00 | Control | Male | 180ms | 3.0T | Moderate |
| 10 | 35.14 | Control | Female | 250ms | 3.0T | Moderate |

**Table 5.** Table of number of subjects (n), mean measurement, and standard deviation (SD) - stratified by healthy control and T21 groups

| | | T21 (mean GA: 32.48) | | | Control (mean GA: 31.88) | | |
|---|---|---|---|---|---|---|---|
| Biometry ID | Biometry name, (unit) | n | Mean | SD | n | Mean | SD |
| 1 | ASBL, (mm) | 24 | 34.45 | 2.20 | 108 | 36.76 | 2.13 |
| 2 | PSBL, (mm) | 24 | 24.10 | 1.40 | 108 | 24.77 | 1.45 |
| 3 | HPL, (mm) | 24 | 24.69 | 2.10 | 108 | 26.02 | 1.85 |
| 4 | VPL, (mm) | 24 | 38.21 | 2.82 | 108 | 40.22 | 2.53 |
| 5 | PAW, (mm) | 24 | 24.76 | 1.71 | 108 | 24.88 | 1.72 |
| 6 | Cho_r, (mm) | 24 | 5.04 | 0.46 | 108 | 5.40 | 0.50 |
| 7 | Cho_l, (mm) | 24 | 5.68 | 0.45 | 108 | 5.94 | 0.53 |
| 8 | NPW, (mm) | 24 | 9.29 | 0.95 | 108 | 9.87 | 1.08 |
| 9 | NB, (mm) | 24 | 7.92 | 0.74 | 108 | 7.41 | 0.69 |
| 10 | PNTh, (mm) | 24 | 7.81 | 1.21 | 108 | 7.14 | 0.88 |
| 11 | OFD, (mm) | 24 | 97.82 | 5.41 | 108 | 101.05 | 6.34 |
| 12 | BPD, (mm) | 24 | 84.43 | 4.17 | 108 | 83.38 | 5.43 |
| 13 | MCh, (mm) | 24 | 79.80 | 4.13 | 108 | 80.36 | 4.26 |
| 14 | OD_R, (mm) | 24 | 17.18 | 0.88 | 108 | 17.35 | 0.98 |
| 15 | OD_L, (mm) | 24 | 17.53 | 0.99 | 108 | 17.80 | 1.07 |
| 16 | IOD, (mm) | 24 | 15.48 | 1.14 | 108 | 15.91 | 1.18 |
| 17 | BOD, (mm) | 24 | 49.95 | 2.64 | 108 | 50.64 | 2.77 |
| 18 | MxW, (mm) | 24 | 23.90 | 1.62 | 108 | 24.25 | 1.67 |
| 19 | MdW, (mm) | 24 | 30.42 | 1.86 | 108 | 30.89 | 2.26 |
| 20 | CBA1, ( $^0$ ) | 24 | 131.20 | 3.76 | 108 | 129.27 | 4.31 |
| 21 | CBA2, ( $^0$ ) | 24 | 116.89 | 5.72 | 108 | 115.39 | 7.50 |
| 22 | FMA, ( $^0$ ) | 24 | 116.28 | 4.56 | 108 | 114.04 | 4.99 |
| 23 | MNMA, ( $^0$ ) | 24 | 17.66 | 2.19 | 108 | 17.91 | 2.27 |
| 24 | IFA, ( $^0$ ) | 24 | 55.16 | 5.22 | 108 | 48.97 | 4.95 |
| 25 | PAH, (mm) | 24 | 3.28 | 0.63 | 108 | 3.00 | 0.58 |
| 26 | ChoH, (mm) | 24 | 3.30 | 0.54 | 108 | 3.22 | 0.59 |
| 27 | MXL, (mm) | 24 | 24.99 | 1.95 | 108 | 26.23 | 1.80 |
| 28 | MDL, (mm) | 24 | 19.02 | 1.68 | 108 | 19.27 | 1.53 |
| 29 | NASO, ($mm^3$) | 24 | 58.42 | 7.18 | 108 | 63.88 | 9.37 |
| 30 | ORO, ($mm^3$) | 24 | 181.41 | 32.81 | 108 | 173.97 | 33.06 |
| 31 | HC, (mm) | 24 | 289.62 | 14.46 | 108 | 296.01 | 18.20 |

**Table 6.** Summary of included datasets for the 3rd trimester T21 and age-matched control cohorts, by GA, MRI protocol, fetal sex and ethnicity

| | | T21 | Control | Total count all, n |
|---|---|---|---|---|
| **Gestational age** | Range | 29.86 - 35.71 | 29.00 - 36.00 | - |
| | Mean (SD) | 32.48 (1.85) | 31.88 (2.01) | - |
| **MRI Protocol, n (%)** | 1.5T / 80ms | 4 (16.6) | 34 (31.5) | 38 |
| | 3T / 180ms | 17 (70.8) | 25 (23.1) | 42 |
| | 3T / 250ms | 3 (0.1) | 49 (45.4) | 52 |
| | **MRI protocol total, n (%)** | **24(100)** | **108 (100)** | 132 |
| **Fetal Sex, n (%)** | Female | 10 (41.7) | 53 (49.1) | 63 |
| | Male | 10 (41.7) | 54 (50.0) | 64 |
| | Unknown | 4 (5.4) | 1 (0.9) | 5 |
| | **Fetal sex total,n (%)** | **24 (100)** | **108 (100)** | 132 |
| **Ethnicity, n (%)** | Asian | 1 (4.2) | 10 (9.3) | 11 |
| | Black | 1 (4.2) | 4 (3.7) | 5 |
| | Other | 0 (0) | 3 (2.8) | 3 |
| | Unknown | 21 (87.5) | 5 (4.6) | 26 |
| | White | 1 (4.2) | 86 (79.6) | 87 |
| | **Ethnicity total, n** (%) | **24 (100)** | **108 (100)** | 132 |

| Variable | Bestfit line equation | SD equation | Mean bestfit method |
|---|---|---|---|
| ASBL | 0.7191 *GA + 13.340 | 0.039 *GA + 0.221 | Linear |
| PSBL | 28.312 + -0.6771 * GA + 0.018 *GA*GA | 0.060 * GA + -0.898 | Quadratic |
| HPL | 0.6207 * GA + 6.230 | 0.068 * GA + -0.843 | Linear |
| VPL | 58.7882 + -2.0538 * GA + 0.0460 * GA*GA | -0.031 * GA + 2.508 | Quadratic |
| PAW | 0.6259 * GA + 4.925 | 0.042 * GA + -0.178 | Linear |
| Cho_r | 0.1199 * GA + 1.575 | 0.019* GA + -0.173 | Linear |
| Cho_l | 0.1438 * GA + 1.359 | 0.049 * GA + -1.150 | Linear |
| NPW | 0.1517 * GA + 5.0364 | -0.022 * GA + 1.71 | Linear |
| NB | 35.307 + -1.940 * GA + 0.032 * GA*GA | 0.008 * GA + 0.188 | Quadratic |
| PNTh | 0.211 * GA + -0.926 | -0.013* GA + 1.085 | Linear |
| OFD | 113.735 + -2.971 * GA + 0.0803 * GA*GA | 0.118 * GA + 0.432 | Quadratic |
| BPD | 1.885 * GA + 23.520 | 0.037 * GA + 2.485 | Linear |
| MCh | 1.410 * GA + 34.461 | 0.119 * GA + -1.101 | Linear |
| OD_R | 0.3607 * GA + 5.856 | 0.011 * GA + 0.242 | Linear |
| OD_L | 0.393 * GA + 5.281 | 0.037 * GA + -0.501 | Linear |
| IOD | 0.2642 * GA + 7.485 | 0.023 * GA + 0.255 | Linear |
| BOD | 1.049 * GA + 17.203 | 0.037 * GA + 0.435 | Linear |
| MxW | 0.609 * GA + 4.849 | 0.081 * GA + -1.470 | Linear |
| MdW | -19.0189+ 2.345 * GA + -0.024 * GA*GA | 0.002 * GA + 1.572 | Quadratic |
| CBA1 | 0.230 * GA + 120.529 | 0.266 * GA + -4.4712 | Linear |
| CBA2 | 1.301 * GA + 79.057 | 0.266 * GA + -4.471 | Linear |
| FMA | 0.091 * GA + 109.872 | 0.140* GA + 0.693 | Linear |
| MNMA | 0.159 * GA + 13.787 | 0.0158 * GA + 1.795 | Linear |
| IFA | 0.253 * GA + 42.129 | 0.206 * GA + -1.307 | Linear |
| PAH | 0.067 * GA + 0.858 | 0.004 * GA + 0.392 | Linear |
| ChoH | 0.0129 * GA + 2.880 | 0.023* GA + -0.126 | Linear |
| MXL | 55.528 + -2.427 * GA + 0.047 * GA*GA | 0.005 * GA + 1.058 | Quadratic |
| MDL | 33.245 + -1.336* GA + 0.028 * GA*GA | 0.0321 * GA + 0.136 | Quadratic |
| NASO | 2.376 * GA + -2.227 | 0.0142 * GA + 8.092 | Linear |
| ORO | 9.157 * GA + -117.920 | 1.535 * GA + -21.413 | Linear |
| HC | 322.684 + -7.907 * GA + 0.221 * GA*GA | -0.145 * GA + 16.694 | Quadratic |

**Table 7.** Best linefit and Standard Deviation (SD) regression equations and method, for each biometric growth chart. GA = Gestational age. Note: all SD regression formulae have linear bestfit method.

| Dependent Variable | Test parameter with covariate | B | Robust Std. Error | t | Sig. | 95% C.I. Lower Bound | 95% C.I. Upper Bound | Partial Eta Squared | Noncent. Parameter | Observed Power |
|---|---|---|---|---|---|---|---|---|---|---|
| Anterior Skull Base Length, ASBL | T21 with GA* | -31.631 | 6.334 | -4.994 | 0.000 | -44.163 | -19.099 | 0.162 | 4.994 | 0.999 |
| Hard Palate Length, HPL | T21 with GA* | -1.682 | 0.329 | -5.107 | 0.000 | -2.334 | -1.030 | 0.168 | 5.107 | 0.999 |
| Velopharyngeal Length, VPL | T21 with GA* | -2.536 | 0.438 | -5.784 | 0.000 | -3.403 | -1.668 | 0.206 | 5.784 | 1.000 |
| Occipitofrontal Diameter, OFD | T21 with GA* | -4.389 | 0.908 | -4.835 | 0.000 | -6.185 | -2.593 | 0.153 | 4.835 | 0.998 |
| Biparietal Diameter, BPD | T21 with GA§ | 0.297 | 0.677 | 0.438 | 0.662§ | -1.043 | 1.636 | 0.001 | 0.438 | 0.072 |
| Inferior Facial Angle, IFA | T21 with GA* | 7.042 | 1.563 | 4.505 | 0.000 | 3.949 | 10.135 | 0.136 | 4.505 | 0.994 |
| Maxillary Length, MXL | T21 with GA* | -1.608 | 0.308 | -5.217 | 0.000 | -2.217 | -0.998 | 0.174 | 5.217 | 0.999 |
| Nasopharyngeal Area, NASO | T21 with GA* | -8.483 | 1.777 | -4.774 | 0.000 | -11.999 | -4.968 | 0.150 | 4.774 | 0.997 |

* = Large effect size
§ = non-significant result and at risk of Type II error
GA = Gestational Age

**Table 8.** ANOVA results for statistically significant biometry with largest effect size in T21 cohort (ABSL, HPL, VPL, OFD, IFA, NASO) and BPD as a non-significant results. Results corrected with robust standard errors.
